# Trade unions and mental health during an employment crisis. Evidence from the UK before and during the COVID-19 pandemic

**DOI:** 10.1101/2023.10.30.23297780

**Authors:** Theocharis Kromydas, Evangelia Demou, Alastair H Leyland, Srinivasa Vittal Katikireddi, Jacques Wels

## Abstract

**Background:** Trade union presence within a workplace could potentially affect employees’ working conditions and in turn health. We assessed the relationship between union (presence and membership) at the individual level and mental health in the context of COVID-19 employment disruptions.

**Methods:** We analysed panel data from Understanding Society collected before and during the COVID-19 pandemic (49,915 observations across 5,988 respondents) to assess the relationship between union presence and membership and a validated epidemiological measure of common mental disorders (CMD), the 12-Item General Health Questionnaire with a score of >/4 indicating probable anxiety/depression, referred to as caseness. A mixed-effect log-linear model assessed effect heterogeneity across time and industries, with average marginal effects (AME) indicating effect differences between groups.

**Findings:** In our sample, 49.1% worked in a unionised workplace, among which 53.8% were union members. Caseness prevalence was higher during the pandemic (25.4%) compared to pre-pandemic (18.4%). Working in a workplace with a trade union was associated with modest protection against CMD risk; (AME_pre-pandemic_:0.010, 95%CI:-0.007; 0.027), (AME_-pandemic_:-0.002, 95%CI:-0.019; 0.016)]. Similarly, for union membership [(AME_pre-pandemic_:0.016, 95%CI:-0.007; 0.039), (AME_pandemic_:-0.010, 95%CI:-0.023; 0.020)]. Industry level heterogeneity exists in the relationship between union presence and membership and mental health.

**Interpretation:** Trade union presence may have a protective effect on workers mental health in periods of crisis, such as during a pandemic. Within unionised workplaces, trade union membership further mitigated the negative effects of the pandemic on mental health. Collective negotiation within workplaces may be protective in periods of uncertainty, benefiting all workers and not only those unionised.

**Funding:** Medical Research Council, Chief Scientist Office, European Research Council, Belgian National Scientific Fund (FNRS).

## Introduction

An emerging amount of evidence has pointed out the relationship between workers’ health and the role of trade unions. Studies have shown that workplace collective negotiation is associated with better workers’ health outcomes and that the lack of such negotiation is often associated with greater vulnerability within the workplace ^1^, the absence of a trade union or staff association within the workplace being associated with both poorer workers’ physical and mental health ^2^. However, the nature of this relationship is complex and the few studies on the topic are contradictory with some demonstrating a negative relationship between trade union membership and physical or mental health ^3^. One of the reasons for conflicting results is the level of collective negotiation that is taken into consideration in explaining population health ^4^. Whilst the general tendency would be to consider union membership – i.e., whether a worker is an actual member of a trade union – as the exposure ^3,5^ other studies have emphasised that the role of trade unions within workplaces goes beyond membership behaviours ^4^. In that sense, unions would also contribute to explain the health of those who are not unionized ^5^. When looking at the workforce, studies have underlined that union presence – that measures whether union representatives are involved in collective bargaining and health and safety committees at a workplace level – is a more relevant distinction because it includes the potential health benefits that affect those in a unionized workplace but who are not members of a trade union.

The distinction is of interest because the relative decline in union membership rates – that is particularly due to structural changes, the rise in non-standard employment and the decline of typically unionized industries ^6^ but also to a shift in personal values ^7,8^ – has contributed to consider unions as not policy relevant anymore. In the United Kingdom, despite having increased slightly over the past few years mainly driven by a surge in female membership, membership rates were around 23.7 percent in 2020 ^9^. By contrast, comparing to union membership levels ^10^, the percentage of employees in the UK covered by a union or workplace council has slightly dropped from 50.2 to 48.2 percent between 1996 and 2018. In other words, the number of workers working in a unionized workplace is more than twice the number of affiliated members ^2^ and one out of two workplaces still has at least one representative trade union ^2^.

The COVID-19 pandemic was a period of high economic disruption, restrictions and changes in employment settings and practices occurred, including an abrupt increase in home working ^11–13^ and the use of temporary unemployment schemes (furlough) ^14^ that had important effect on population health, and particularly mental health ^15^. Whilst the relationship between change in employment status and work pattern and workers’ mental health have been documented throughout this period ^14,16^, less is known about the way workplace characteristics – and, particularly, trade union presence – may have affected mental health. Trade unions played a crucial role during such a period, advocating for worker safety, protection of jobs, and fair compensation for those affected by the economic fallout of the pandemic ^17,18^. The COVID-19 has been seen as an occupational disease and unions have worked to advocate and protect the workforce in key sectors of activity ^19,20^. Whilst social dialogue between social partners and the state has been slow in delivering policies or action during the pandemic in the UK ^21^, the role of trade unions was pivotal at company-level where collective negotiation, participation in Safety and Health Committees^22^ and greater job stability ^23^ might have had a protective role on working conditions. A report by the Trade Union Congress, highlighted how unions remained active during the pandemic and increased the number of health and safety representatives in an effort to reduce the impacts of the pandemic on workers ^24^.

No study has examined such a trend during the pandemic and pre-pandemic studies are few ^2–4,25^. To better assess such a relationship, this study focused on workers’ mental health changes observed between a period of a health crisis such as the COVID-19 pandemic where population and workers mental health deteriorated and a period of time before pandemic started^15,26–28^. The primary research objective of the study is to address whether workers in unionized workplaces experienced the same changes in mental health during the pandemic compared to workers in non-unionized workers. Additionally, we investigate whether any changes was observed by sector of activity and whether the same patterns are revealed when union membership is used as an exposure instead of trade union presence. By addressing these, the study aims to evidence the nature of the relationship between trade unions and workers’ mental health, to establish whether trade union presence is a key variable associated with workers’ health and, more fundamentally, to question whether diminishing trade union influence and bargaining power might translate into a decline in the workforce’s mental health.

## Data and methods

### Understanding Society (USoc)

We used data from Understanding Society waves 9, 10 and 11 (baseline sample) and all nine COVID-19 sweeps collected throughout 2020 and 2021: sweep 1 (April 2020), sweep 2 (May 2020), sweep 3 (June 2020), sweep 4 (July 2020), sweep 5 (September 2020), sweep 6 (November 2020), sweep 7 (January 2021) sweep 8 (March 2021) and sweep 9 (September 2021). The analytical sample is restricted to respondents who participated in wave 9, were employed at that time (excluding self-employed workers – but not those who combined employment and self-employment) and participated in, at least, one of the nine COVID-19 sweeps (see <u>supplementary file S1</u>).

### Outcome variables

GHQ-12 caseness was our outcome of interest. It is derived from the 12-Item General Health Questionnaire (GHQ-12) based on the answers to 12 questions that assess severity of mental problems over the past few weeks ^29^.The GHQ-12 variable is a validated screening tool for identifying common mental disorders (i.e. probable anxiety and/or depression) and it was used on a binary basis (GHQ-caseness or not) based on cut-off score of 4 ^30^.

### Exposure variables

We used two exposure variables and replicated the analyses for each. *Union presence* was defined by participants yes-no response to the following question: "Is there a trade union, or a similar body such as a staff association, recognised by your management for negotiating pay or conditions for the people doing your sort of job in your workplace?” We also used Usoc variable on *union membership* that applies only to respondents in workplaces where there is a union or staff association and distinguished between members and non-members. Data on union presence and union membership for the employed workforce were only collected in pre-pandemic waves 2, 4, 6, 8 and 10. We carried forward union presence and union membership variables’ values for wave 9 and 11 using available data in waves 8 and 10, respectively. Then we carried all data forward to all COVID-19 sweeps. This data generation technique was based on the relative stability of occupation and industry across time ^27,31^, and by testing changes in union presence and membership over time in the pre-pandemic period. The latter showed very high overall stability across waves, which supports our assumption that carrying data forward will be unlikely to bias our results (see <u>Supplementary file S1</u>, Table S1.1 and S1.2).

### Covariates

The model controls for a set of socio-economic and demographic confounders. *Gender*(female and male); *Age* and a quadratic function of age; *Ethnicity*: (non-white, white); *Country* of residence: (Scotland, Wales, Northern Ireland and England); Highest level of *education* achieved (A-level, GCSE, other, none or university degree);^28^ (*Company size*: 1 to 34, 25 to 199, more than 200); Self-reported *financial situation* was used as a dummy variable distinguishing those living comfortably, doing alright combined- (coded as 0, ref.) and those finding it very difficult, difficult to cope or just about getting by; and *Heath status*, (reporting a long-standing illness or impairment.: yes, no). Variables were measured at each time point apart from self-reported financial situation, health status, company size, industry and level of education where data were carried forward from the closest available wave. The health status reflects respondents’ report of a long-standing (more than 12 months) physical or mental impairment, illness or disability to reflect baseline health. Moreover, data for the self-reported financial situation were carried one COVID sweep backwards when non-available in the current sweep or previous ones. We included dummy variables for waves of participation to account for secular effects across time and we further include an *Industry* variable including 15 levels for the Standard Industry Classification (SIC) as used in ^27^ as an effect modifier.

### Statistical analysis

We used a mixed-effects analysis with a log-linear link function (for binary outcomes) accounting for repeated observations over time on the individual level and adjusting for serial correlation of observations within individual values using the cluster sandwich estimator, which allows for intragroup correlation in standard errors. We estimated unadjusted and adjusted models using both odds ratios (reported in the <u>Supplementary file S2</u>) and predicted probabilities (predicted marginal means and average marginal effects). The unadjusted model (model 1) regresses our binary outcome variable over our exposures by using a 2-way interaction between trade union presence/membership and a binary variable that shows pre-pandemic (waves, 9,10 and 11) and pandemic periods (all COVID-19 sweeps). The adjusted model (model 2) includes the same 2-way interaction but controls for the full set of covariates. Finally, we included an industry classification term creating a 3-way interaction, which was our effect modifier in model 3. Analyses were replicated using USoc-provided sampling weights with no major difference across estimates. Additionally, we calculated Inverse Probability Weights (IPW) for our outcome missingness separately for each analytical sample used and wave/sweep based on basic demographics (age, sex, UK country of residence, ethnicity and trade union presence or trade union membership). This results to each individual assigned a weight that is the inverse of their probability of being a complete case. The weights used for our final estimates were the product of the Usoc-provided sampling weights and the IPW we calculated for outcome missingness.

To better interpret the estimates flowing from the logistic regression models, we converted odds ratios to probabilities (additive instead of multiplicative effects) using the Stata margins command focusing on the fixed part of the mixed-effects regression equation. We have estimated both the marginal means (MM) and average marginal effects (AME).

### Sensitivity Analyses

We conducted three types of sensitivity analyses by replicating our analytical methodology over two sub-samples. First, we excluded trade union members to check the specific relationship between union presence and GHQ-12 caseness for the non-unionized workforce and then we excluded publicly owned workplaces, where the proportion of unionized workplaces in our data is larger compared to private-owned workplaces. Finally, we run a linear model using a variable that measures mental health on broader scale (GHQ-36) as our outcome variable. We identify no differences in the direction of the effect compared to our binary logistic model.

## Results

The study included 49,915 observations among 5,988 participants (total sample) and 27,971 observations among 3,341 participants after restricting the sample to those reporting union presence (Supplementary file S1).

**Table 1** shows that, among the respondents who were in employment at data collection time, 50.8 percent reported having a trade union within their workplace (weighted data) before the start of the pandemic against 48.5 percent during the pandemic. In other words, union presence applied to about half of the sample. When looking at the population working in a unionized workplace, we observe that trade union membership is 57.4 percent in pre-pandemic waves and 52.5 percent during the pandemic. More than half of the workforce in unionized workplaces is a trade union or staff association member.

**Table 1.**
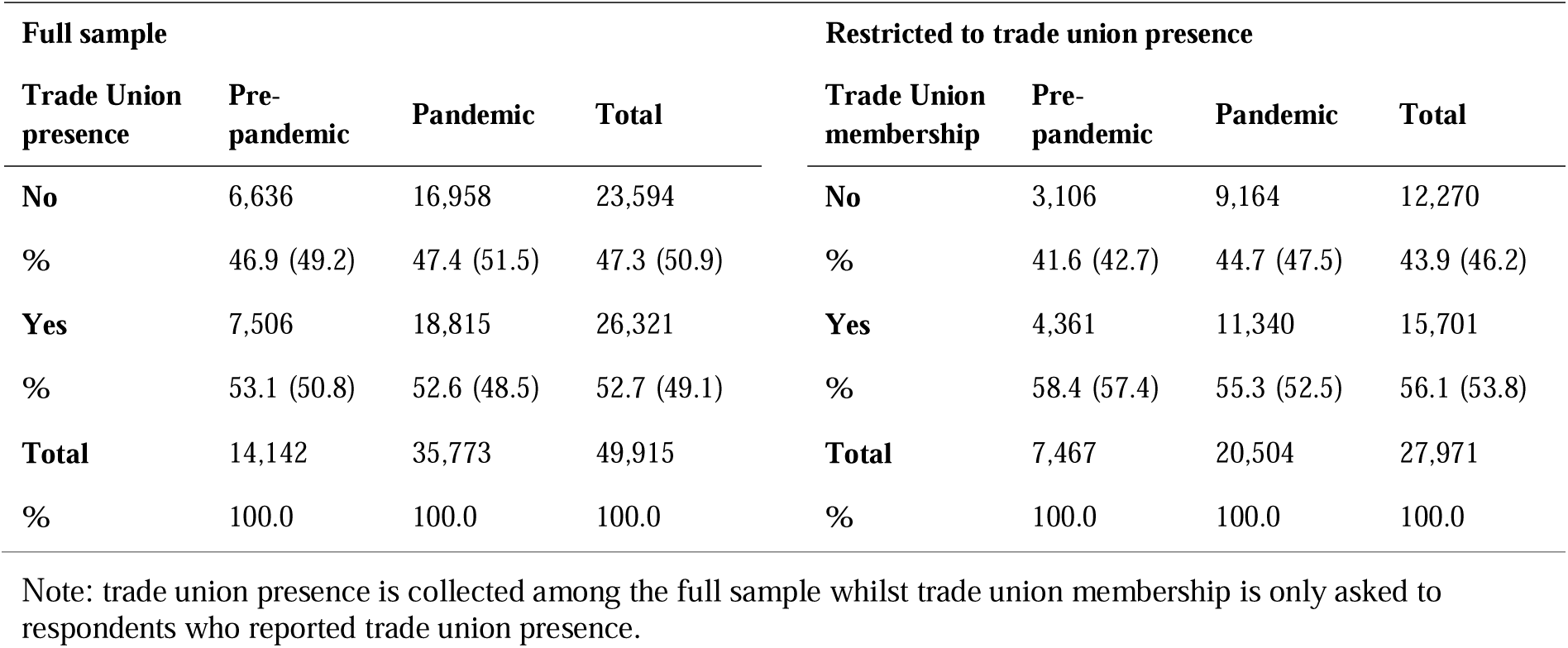
Trade union presence and membership in pre-pandemic and pandemic waves (unweighted and weighted sample (in brackets))

| Full sample |  |  |  | Restricted to trade union presence |  |  |  |
| --- | --- | --- | --- | --- | --- | --- | --- |
| Trade Union presence | Pre-pandemic | Pandemic | Total | Trade Union membership | Pre-pandemic | Pandemic | Total |
| **No** | 6,636 | 16,958 | 23,594 | **No** | 3,106 | 9,164 | 12,270 |
| **%** | 46.9 (49.2) | 47.4 (51.5) | 47.3 (50.9) | **%** | 41.6 (42.7) | 44.7 (47.5) | 43.9 (46.2) |
| **Yes** | 7,506 | 18,815 | 26,321 | **Yes** | 4,361 | 11,340 | 15,701 |
| **%** | 53.1 (50.8) | 52.6 (48.5) | 52.7 (49.1) | **%** | 58.4 (57.4) | 55.3 (52.5) | 56.1 (53.8) |
| **Total** | 14,142 | 35,773 | 49,915 | **Total** | 7,467 | 20,504 | 27,971 |
| **%** | 100.0 | 100.0 | 100.0 | **%** | 100.0 | 100.0 | 100.0 |

Note: trade union presence is collected among the full sample whilst trade union membership is only asked to respondents who reported trade union presence.

Further descriptive statistics on sample composition are shown in <u>Supplementary file S3</u> (Tables S3.1, S3.2). For our analytical sample including both unionised and non-unionised workplaces, GHQ-12 caseness was 17.7 percent before the pandemic and 23.8 percent during the pandemic (respectively 18.4 and 25.4 percent when data are weighted). When data are restricted to workplaces where there is a trade union present, GHQ-12 caseness is 18.6 and 24.4 percent (19.4 and 25.3 percent after weighting), pre and during the pandemic respectively (Table S3.1).

We also observe a difference in industry representation depending on whether the sample is restricted to workplaces where a trade union is present. For instance, whilst the composition of workers in the public administration and defence industry is 10.1 and 7.2 percent in the full sample in pre-pandemic and pandemic waves, these reach 17.0 and 11.2 when the sample is restricted to union presence. The same pattern can be observed in other industries, which are heavily represented by the public sector. For instance, sample composition for Education are

9.3 and 8.3 percent higher in the restricted sample. Similarly, for Human Health and Social Work Activities the equivalent compositional percentages were 5 and 4.6 (Table S3.2). This indicates that union presence in these industries is particularly salient.

In **Table 2,** the AMEs refer to the difference in the prevalence of GHQ-12 caseness between respondents in unionised and non-unionised workplaces and were estimated separately for the pre- and pandemic periods for both samples used. Negative AMEs denote higher prevalence for respondents in non-unionised workplaces (2a) and trade union non-members (2b). In the adjusted Model 1 MMs show that the average GHQ-12 caseness prevalence in unionised workplaces is slightly higher pre-pandemic (0.121) compared to non-unionised workplaces (0.111) but this trend reverses in the pandemic period (i.e., 0.148 vs 0.150, respectively). This is reflected by the positive AME sign pre-pandemic (0.010, 95%CI: -0.007; 0.027) and the negative sign during the pandemic period (-0.002, 95%CI: -0.019; 0.016). Undoubtedly, on average, there has been a deterioration of mental health of the working population during the pandemic compared to before. However, results from model 1 indicate trade union presence mitigated that negative change, although estimates are imprecise.

**Table 2:** Adjusted Models (Model 2 and Model 3) Marginal Means (MM) and Average Marginal Effects (AVE) for Trade Union Presence and membership as exposures.

|  |  | Full sample |  |  |  |  |  |  | Restricted to trade union presence |  |  |  |  |  |
| --- | --- | --- | --- | --- | --- | --- | --- | --- | --- | --- | --- | --- | --- | --- |
|  | Time Period | Trade Union | MM | 95 % CI |  | AME | 95 % CI |  | MM | 95 % CI |  | AME | 95 % CI |  |
|  |  |  |  | Lower | Higher |  | Lower | Higher |  | Lower | Higher |  | Lower | Higher |
| **Model 1** |  |  |  |  |  |  |  |  |  |  |  |  |  |  |
| Total sample (TS) | Pre-pandemic | No | 0.111 | 0.076 | 0.439 |  |  |  | 0.119 | 0.096 | 0.142 |  |  |  |
|  | Pre-pandemic | Yes | 0.121 | 0.048 | 0.276 | 0.010 | -0.007 | 0.027 | 0.135 | 0.111 | 0.158 | 0.016 | -0.007 | 0.039 |
|  | pandemic | No | 0.150 | 0.135 | 0.283 |  |  |  | 0.155 | 0.138 | 0.172 |  |  |  |
|  | pandemic | Yes | 0.148 | 0.064 | 0.233 | -0.002 | -0.019 | 0.016 | 0.154 | 0.137 | 0.170 | -0.001 | -0.023 | 0.020 |
| **Model 2 Industries** |  |  |  |  |  |  |  |  |  |  |  |  |  |  |
| Mining, Energy and Water Supply (MEWS) | Pre-pandemic | No | 0.258 | 0.076 | 0.439 |  |  |  | 0.141 | -0.032 | 0.313 |  |  |  |
|  | Pre-pandemic | Yes | 0.162 | 0.048 | 0.276 | -0.096 | -0.307 | 0.116 | 0.197 | 0.044 | 0.350 | 0.056 | -0.174 | 0.286 |
|  | pandemic | No | 0.209 | 0.135 | 0.283 |  |  |  | 0.157 | 0.018 | 0.295 |  |  |  |
|  | pandemic | Yes | 0.148 | 0.064 | 0.233 | -0.061 | -0.172 | 0.050 | 0.146 | 0.067 | 0.226 | -0.010 | -0.168 | 0.148 |
| Manufacturing (MAN) | Pre-pandemic | No | 0.101 | 0.070 | 0.133 |  |  |  | 0.043 | -0.003 | 0.088 |  |  |  |
|  | Pre-pandemic | Yes | 0.070 | 0.031 | 0.110 | -0.031 | -0.077 | 0.015 | 0.107 | 0.045 | 0.169 | 0.064 | -0.008 | 0.137 |
|  | pandemic | No | 0.146 | 0.110 | 0.182 |  |  |  | 0.099 | 0.040 | 0.158 |  |  |  |
|  | pandemic | Yes | 0.098 | 0.057 | 0.138 | -0.048 | -0.103 | 0.007 | 0.116 | 0.059 | 0.173 | 0.017 | -0.064 | 0.098 |
| Construction (CON) | Pre-pandemic | No | 0.085 | 0.044 | 0.127 |  |  |  | 0.221 | 0.042 | 0.400 |  |  |  |
|  | Pre-pandemic | Yes | 0.168 | 0.051 | 0.285 | 0.083 | -0.038 | 0.204 | 0.156 | -0.007 | 0.320 | -0.065 | -0.305 | 0.175 |
|  | pandemic | No | 0.130 | 0.090 | 0.170 |  |  |  | 0.128 | 0.048 | 0.209 |  |  |  |
|  | pandemic | Yes | 0.140 | 0.071 | 0.208 | 0.010 | -0.069 | 0.088 | 0.153 | 0.058 | 0.248 | 0.025 | -0.099 | 0.148 |
| Wholesale and Retail Trade Motor Repair (WRTMR) | Pre-pandemic | No | 0.123 | 0.084 | 0.162 |  |  |  | 0.156 | 0.073 | 0.238 |  |  |  |
|  | Pre-pandemic | Yes | 0.131 | 0.085 | 0.176 | 0.008 | -0.048 | 0.064 | 0.132 | 0.070 | 0.194 | -0.024 | -0.128 | 0.081 |
|  | pandemic | No | 0.167 | 0.129 | 0.205 |  |  |  | 0.168 | 0.108 | 0.227 |  |  |  |
|  | pandemic | Yes | 0.137 | 0.097 | 0.178 | -0.030 | -0.084 | 0.025 | 0.140 | 0.085 | 0.195 | -0.027 | -0.106 | 0.052 |
| Transportation and Storage (TS) | Pre-pandemic | No | 0.111 | 0.051 | 0.172 |  |  |  | 0.064 | 0.006 | 0.122 |  |  |  |
|  | Pre-pandemic | Yes | 0.096 | 0.052 | 0.141 | -0.015 | -0.085 | 0.055 | 0.130 | 0.064 | 0.196 | 0.066 | -0.019 | 0.151 |
|  | pandemic | No | 0.104 | 0.056 | 0.153 |  |  |  | 0.133 | 0.051 | 0.215 |  |  |  |
|  | pandemic | Yes | 0.155 | 0.104 | 0.206 | 0.051 | -0.019 | 0.121 | 0.178 | 0.115 | 0.241 | 0.045 | -0.057 | 0.148 |
| Accommodation and Food Services (AF) | Pre-pandemic | No | 0.172 | 0.102 | 0.242 |  |  |  | 0.107 | 0.001 | 0.214 |  |  |  |
|  | Pre-pandemic | Yes | 0.070 | 0.005 | 0.135 | -0.102 | -0.196 | -0.008 | 0.008 | -0.008 | 0.024 | -0.100 | -0.208 | 0.008 |
|  | pandemic | No | 0.177 | 0.117 | 0.236 |  |  |  | 0.183 | 0.098 | 0.268 |  |  |  |
|  | pandemic | Yes | 0.174 | 0.090 | 0.258 | -0.003 | -0.105 | 0.100 | 0.157 | 0.028 | 0.287 | -0.026 | -0.181 | 0.129 |
| Information and Communication (IC) | Pre-pandemic | No | 0.090 | 0.041 | 0.138 |  |  |  | 0.175 | 0.027 | 0.323 |  |  |  |
|  | Pre-pandemic | Yes | 0.233 | 0.078 | 0.388 | 0.143 | -0.016 | 0.303 | 0.266 | 0.047 | 0.486 | 0.092 | -0.167 | 0.350 |
|  | pandemic | No | 0.172 | 0.132 | 0.211 |  |  |  | 0.201 | 0.108 | 0.294 |  |  |  |
|  | pandemic | Yes | 0.144 | 0.086 | 0.202 | -0.028 | -0.098 | 0.041 | 0.106 | 0.048 | 0.165 | -0.095 | -0.204 | 0.015 |
| Financial and Insurance Activities (FI) | Pre-pandemic | No | 0.158 | 0.097 | 0.220 |  |  |  | 0.062 | 0.019 | 0.106 |  |  |  |
|  | Pre-pandemic | Yes | 0.104 | 0.045 | 0.162 | -0.055 | -0.135 | 0.026 | 0.171 | 0.036 | 0.306 | 0.109 | -0.030 | 0.247 |
|  | pandemic | No | 0.122 | 0.087 | 0.158 |  |  |  | 0.113 | 0.063 | 0.164 |  |  |  |
|  | pandemic | Yes | 0.103 | 0.061 | 0.145 | -0.019 | -0.073 | 0.034 | 0.077 | 0.023 | 0.132 | -0.036 | -0.110 | 0.038 |
| Real Estate Activities (RE) | Pre-pandemic | No | 0.100 | 0.014 | 0.186 |  |  |  | 0.277 | 0.104 | 0.450 |  |  |  |
|  | Pre-pandemic | Yes | 0.316 | 0.163 | 0.468 | 0.215 | 0.035 | 0.395 | 0.367 | 0.108 | 0.625 | 0.090 | -0.214 | 0.394 |
|  | pandemic | No | 0.168 | 0.079 | 0.257 |  |  |  | 0.138 | 0.013 | 0.263 |  |  |  |
|  | pandemic | Yes | 0.100 | 0.017 | 0.183 | -0.068 | -0.191 | 0.055 | 0.070 | 0.018 | 0.123 | -0.068 | -0.201 | 0.066 |
| Professional Scientific & Technical (PST) | Pre-pandemic | No | 0.103 | 0.072 | 0.133 |  |  |  | 0.114 | 0.033 | 0.196 |  |  |  |
|  | Pre-pandemic | Yes | 0.106 | 0.052 | 0.160 | 0.004 | -0.055 | 0.062 | 0.114 | 0.032 | 0.196 | 0.000 | -0.113 | 0.112 |
|  | pandemic | No | 0.153 | 0.123 | 0.184 |  |  |  | 0.197 | 0.137 | 0.257 |  |  |  |
|  | pandemic | Yes | 0.165 | 0.126 | 0.204 | 0.011 | -0.037 | 0.060 | 0.157 | 0.109 | 0.206 | -0.040 | -0.116 | 0.036 |
| Administrative and Support Services (AS) | Pre-pandemic | No | 0.075 | 0.034 | 0.115 |  |  |  | 0.042 | -0.010 | 0.094 |  |  |  |
|  | Pre-pandemic | Yes | 0.089 | 0.010 | 0.169 | 0.015 | -0.071 | 0.100 | 0.220 | -0.009 | 0.450 | 0.179 | -0.049 | 0.407 |
|  | pandemic | No | 0.124 | 0.083 | 0.165 |  |  |  | 0.146 | 0.092 | 0.200 |  |  |  |
|  | pandemic | Yes | 0.133 | 0.092 | 0.174 | 0.009 | -0.048 | 0.066 | 0.104 | 0.058 | 0.149 | -0.042 | -0.112 | 0.028 |
| Public Administration and Defence (PAD) | Pre-pandemic | No | 0.058 | -0.002 | 0.118 |  |  |  | 0.116 | 0.073 | 0.160 |  |  |  |
|  | Pre-pandemic | Yes | 0.120 | 0.091 | 0.150 | 0.062 | -0.003 | 0.127 | 0.134 | 0.095 | 0.173 | 0.018 | -0.035 | 0.071 |
|  | pandemic | No | 0.126 | 0.074 | 0.178 |  |  |  | 0.137 | 0.103 | 0.172 |  |  |  |
|  | pandemic | Yes | 0.153 | 0.127 | 0.179 | 0.027 | -0.030 | 0.085 | 0.170 | 0.134 | 0.207 | 0.033 | -0.016 | 0.081 |
| Education (EDU) | Pre-pandemic | No | 0.067 | 0.033 | 0.102 |  |  |  | 0.125 | 0.085 | 0.166 |  |  |  |
|  | Pre-pandemic | Yes | 0.118 | 0.092 | 0.143 | 0.051 | 0.012 | 0.089 | 0.125 | 0.091 | 0.159 | 0.000 | -0.046 | 0.046 |
|  | pandemic | No | 0.162 | 0.122 | 0.202 |  |  |  | 0.159 | 0.126 | 0.192 |  |  |  |
|  | pandemic | Yes | 0.155 | 0.133 | 0.177 | -0.007 | -0.051 | 0.037 | 0.163 | 0.136 | 0.190 | 0.004 | -0.037 | 0.045 |
| Human Health and Social Work Activities (HHSW) | Pre-pandemic | No | 0.119 | 0.086 | 0.152 |  |  |  | 0.143 | 0.100 | 0.186 |  |  |  |
|  | Pre-pandemic | Yes | 0.129 | 0.103 | 0.155 | 0.010 | -0.027 | 0.047 | 0.129 | 0.096 | 0.163 | -0.013 | -0.061 | 0.035 |
|  | pandemic | No | 0.140 | 0.110 | 0.170 |  |  |  | 0.169 | 0.137 | 0.202 |  |  |  |
|  | pandemic | Yes | 0.162 | 0.139 | 0.185 | 0.022 | -0.015 | 0.059 | 0.162 | 0.135 | 0.190 | -0.007 | -0.048 | 0.034 |
| Other Services (OTH) | Pre-pandemic | No | 0.134 | 0.075 | 0.193 |  |  |  | 0.044 | 0.010 | 0.078 |  |  |  |
|  | Pre-pandemic | Yes | 0.105 | 0.047 | 0.163 | -0.029 | -0.110 | 0.052 | 0.241 | 0.107 | 0.375 | 0.197 | 0.060 | 0.333 |
|  | pandemic | No | 0.137 | 0.108 | 0.165 |  |  |  | 0.112 | 0.067 | 0.157 |  |  |  |
|  | pandemic | Yes | 0.124 | 0.091 | 0.156 | -0.013 | -0.055 | 0.029 | 0.144 | 0.102 | 0.186 | 0.032 | -0.029 | 0.093 |

We replicated model 1 using union membership as an exposure variable within a sample that only includes respondents who worked workplaces with unions or staff associations (**Table 2)**. An almost identical pattern was observed as for union presence but with even wider confidence intervals. The pre-pandemic AME was 0.016 (95%CI: -0.007; 0.039) and the pandemic AME was -0.001 (95%CI: -0.023; 0.020).

These estimates are summarised in **figure 1** which shows the marginal means for trade union presence (within the full working population) and trade union membership (within the full population working in a unionized workplace) before the outbreak of the COVID-19 pandemic and during the pandemic. What can be observed is that the probability of GHQ-12 caseness increased during the pandemic for all workers irrespective of whether they work in a unionized workplace or were a member of a trade union. However, the increase for those working in a unionized workplace has been sharper: the probability of GHQ-12 caseness for those in workplaces without union presence was 11.12 percent [95% CI:9.42;12.83] prior to the pandemic against 14.99 percent [95% CI:13.68; 16.30] during the pandemic, with non-overlapping confidence intervals. By contrast, for the unionised workplaces the equivalent difference shows confidence intervals that overlap. The probability of GHQ-12 caseness was 12.14 percent [95% CI:10.45; 13.83] prior to the pandemic and 14.82 percent [95% CI:13.51; 16.14] during the pandemic. This implies that union presence in workplaces has an apparent cushioning effect for the mental health of workers compared to union absence. When examining union membership as the exposure, differences across predictive margins are similar but CIs become even wider, principally because the sample was restricted to those in unionized workplaces. Our estimates show a difference between members and non-members before the start of the pandemic with a probability of 11.87 percent [95% CI: 9.57; 14.17] for the those who are not trade union members and 13.49 percent [95% CI: 11.14; 15.84] for those who are. This difference seems to have reversed during the pandemic with respectively 15.50 percent [95% CI: 13.76; 17.23] and 15.36 percent [95% CI: 13.74; 16.97], indicating a potential cushioning effect of union membership, although CIs are wide and overlap.

**Figure 1.**
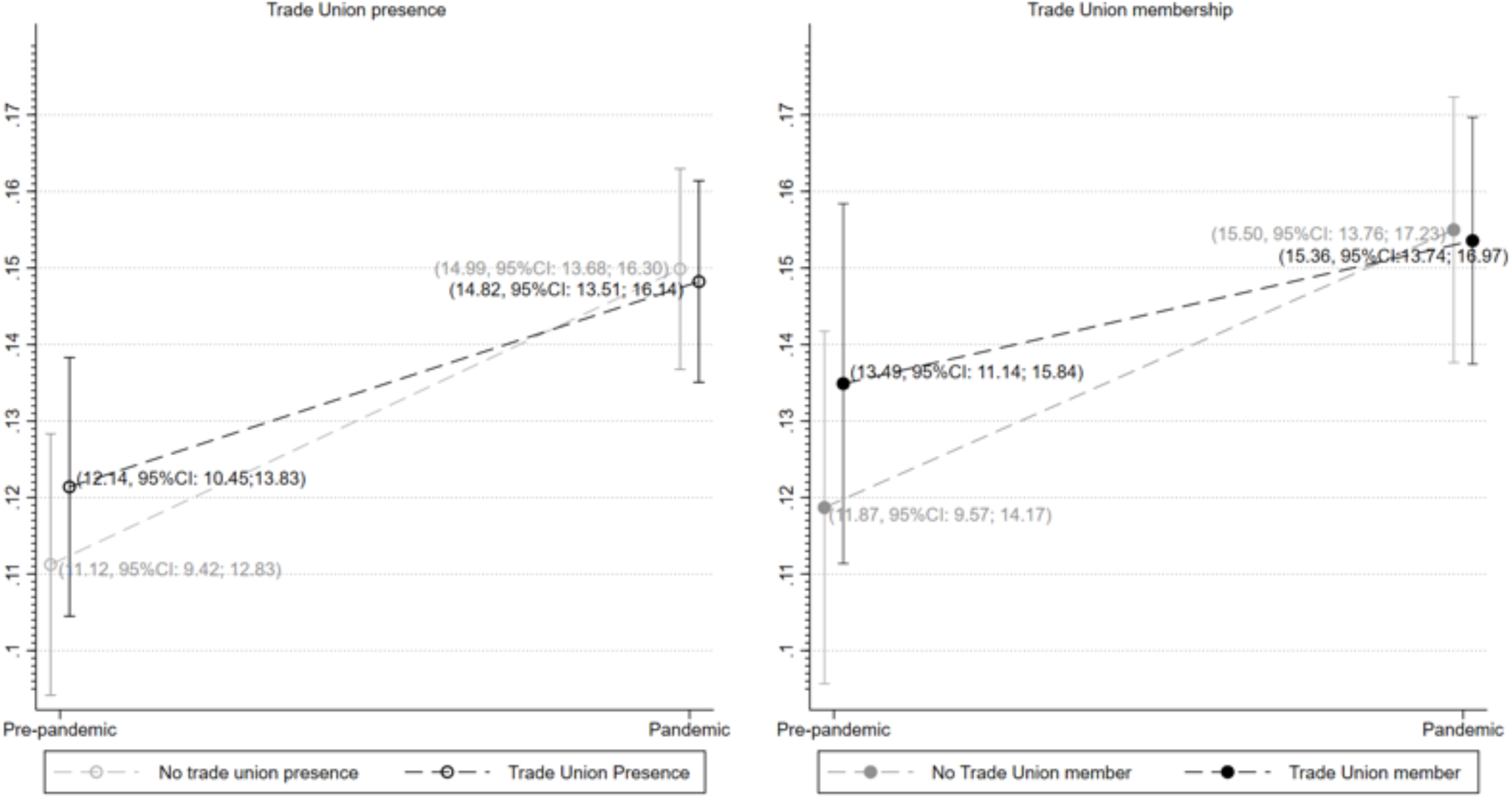
Marginal means of GHQ-12 caseness in pre-pandemic and pandemic periods by union presence and union membership.

What is observed above is an average result that does not consider the potential differential trends that could be observed across industries. Once industry classification is added as term of the interaction effect (Model 2), we observe effect magnitude and direction variations of trade union presence (and membership) by industry, although with very broad confidence intervals that do not allow us drawing meaningful conclusions in most groups due to sample limitations. In only just three (Accommodation and Food services [AME:10.2% CI: -19.6; - 0.1], Real Estate Activities [AME:21.5% CI: 3.5; 39.5] and Education [AME:5.1% CI: 1.2; 8.9]) CIs do not cross zero and also this stands only for pre-pandemic period (Table 2). Thus, we are unable to draw statistically robust conclusions regarding the comparison of the AMEs between the pre-pandemic and pandemic periods. When we use trade union membership as an exposure, confidence intervals are too wide and cross zero in all industries.

These estimates are summarized in figure 2. Full estimates (in odds ratios) including exposure and covariates derived from the logistic regression models are shown in <u>supplementary file S3</u> for both union presence and membership including the unadjusted and adjusted versions of all models.

**Figure 2.**
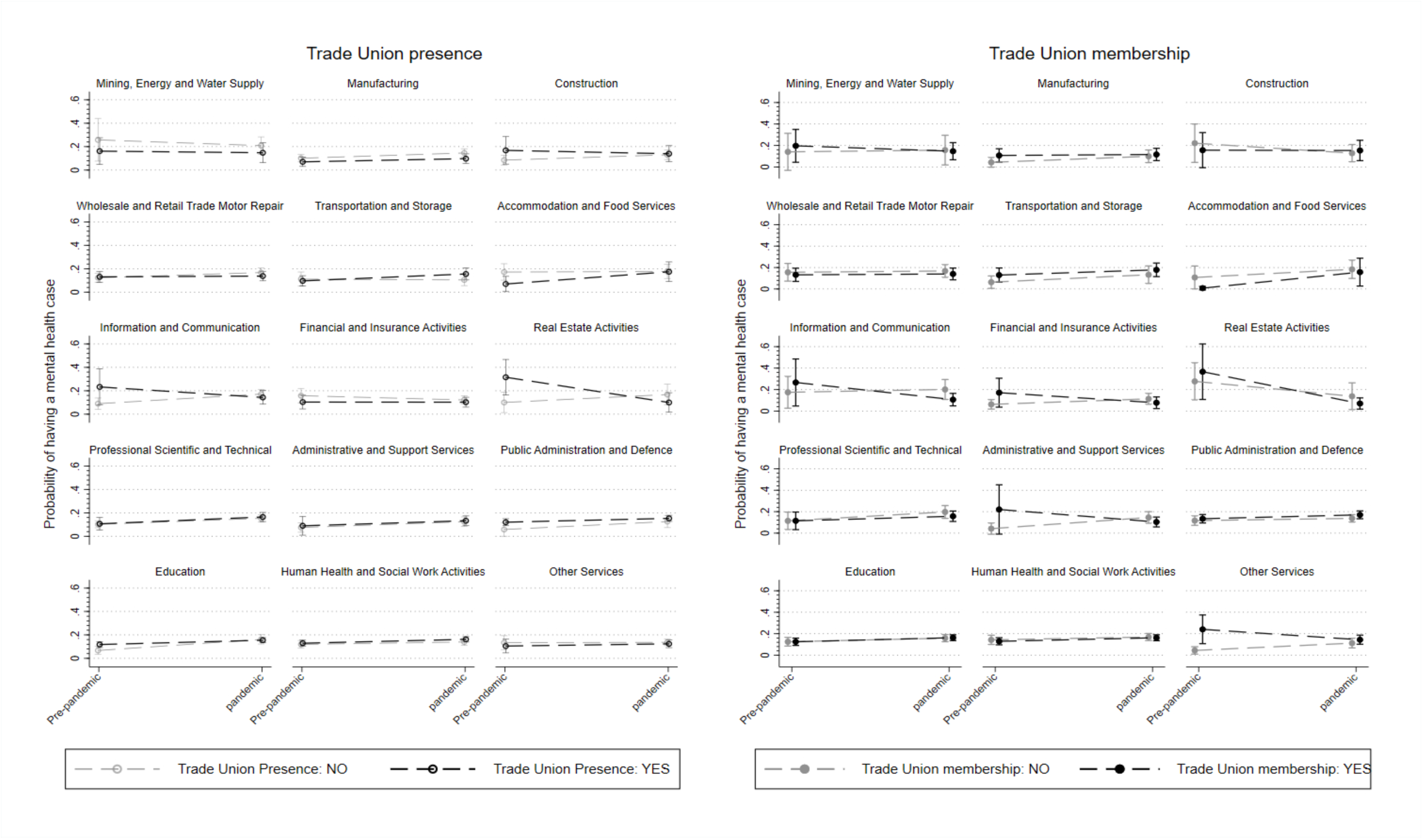
Marginal means of GHQ-12 caseness in pre-pandemic and pandemic waves by union presence and union membership stratified by industry.

Results from sensitivity analyses for (1) union presence excluding public companies, (2) restricted sample to non-unionized workers and (3) using GHQ-36 as a numeric outcome are shown in <u>supplementary file S4</u>. These sensitivity analyses do not show major changes in points estimates for the association between union presence and GHQ-12 caseness indicating that publicly owned companies’ prevalence in some specific industries do not explain differences in GHQ-12 caseness and also that union membership is not a driver of lower GHQ-12 caseness in companies where a trade union is present. Similarly using GHQ-36 instead of GHQ-12 caseness does not affect the direction of the point estimates.

## Discussion

Two main findings flow from this study. First, we demonstrate that whilst the whole workforce’s mental health has declined after the start of the COVID-19 pandemic, the increase in GHQ-12 caseness was proportionally higher among non-unionised compared to unionised workplaces. The same trend is observed when looking at union membership (after restricting the sample to workers working in a unionized workplace only) but results are not robust to draw inferences. Second, heterogeneity exists across industries and there is no consistent pattern on the effect of trade unions on mental health across time in any.

The long-term decline in trade union membership in the United Kingdom poorly reflects the role trade unions continue to play within British companies. With about fifty per cent of the workforce working in a unionised company and roughly a quarter belonging to a trade union, the implications of trade union presence on workers’ health are significant and, yet few studies have investigated such a relationship. Whilst the role of employment characteristics in explaining population health is well documented ^32^, studies on the association between collective negotiation and workers’ health are very few. Previous studies mainly focus on three types of approaches. A first set of studies, that is the most common among the literature pays attention to the relationship between union membership and health using cross-sectional, cross-sectorial or macro-level data. For instance, it has been recently demonstrated using comparative cross-sectional data that health inequalities are high when unions only represent part of the workforce but low when a high proportion of the workforce is unionized ^5^. Similarly, high country trade union density (i.e., calculated based on union membership rates) is associated with lower depressive symptoms among the workforce ^33^. The same type of analyses was also made looking at differences across sectors of activity based on union densities ^34,35^. A second type of studies takes a collective approach by focusing on the negotiating process within health and safety committees that exist in most OECD countries. Those committees are set up to negotiate within the workplace (but also at sector level) working conditions and safety matters and involve trade unions (when the company has trade union representatives) or workers’ representatives. For instance, using Korean cross-sectional data, it was shown that health and safety committees reduce work accidents but seem to be more effective in unionized workplaces ^36^. By contrast, Bryson has shown for the UK that union representation within health and safety committees is linked with lower health and safety risks compared with non-unionized workplaces ^22^. More recently, a study has linked company-based social dialogue quality and workers’ health as perceived by trade unions representatives during the pandemic ^37^. A third type of studies has very recently focused on the individual relationship between union membership – and, to a lesser extent, presence – and health mainly using individual longitudinal data in the US and the UK. These studies have demonstrated contradictory results that show either a positive ^38,39^ or a negative relationship ^3^ mainly due to the nature of the variables used to capture trade union presence or membership and the control variables included in the models. A few other studies have focused on the benefits of using a longitudinal approach to assess the association between union membership and wages ^40^ or job satisfaction ^41^ but such a perspective is still rare when looking at health, and particularly mental health.

Yet, most of these approaches have an individualistic view on unions assuming that any potential benefits gained apply only to members. As many studies distinguish union members and non-union members, the indirect role of union presence in company-based collective negotiations or health and safety committees in protecting those who are not unionized has been ignored ^4^. By looking at both union presence and union membership and using individual longitudinal data, this study is one of the few that assesses the indirect benefits of trade union presence in the workplace. This study also focuses on the difference between pre-pandemic and pandemic time periods. Whilst the relationship between changes in employment settings and workers’ health has been largely documented throughout the pandemic ^12,42–44^, we do not know any quantitative research published on the possible role of tr unions in providing cushioning to the deterioration of workers’ mental health during the COVID-19 pandemic and the number of qualitative studies on this matter remains low, mostly theoretical and address the mechanisms of protection without estimating their prevalence for the entire workforce ^45^.

This study has limitations, which can open the way for further research on collective negotiation and workers’ health. A first limitation is about the nature of the dataset. Understanding Society did not collect information on trade union presence and membership and industry classification during the pandemic and information had to be carried forward from pre-pandemic waves. Similarly, this stands for some of the covariates used. Industry or even company changes were not likely to occur to a large extent during the pandemic, however they were not impossible, and this study overlooks these potential changes due to data limitations. A second limitation is that the study did not account for changes in employment status throughout the pandemic. Whilst respondents who moved to furlough (i.e., the UK job retention scheme) are included in the sample, we did not account for those who became unemployed or were made redundant during the pandemic. A third limitation is that this study did not include information on home-working due to lack of data availability over the selected waves. Whilst trade unions were reluctant to implement home-working policies before the pandemic ^46^ and home-working propensities were lower within the unionized workforce ^46^, the specific effect of trade unions for those working from home is still to be investigated. A fourth limitation is about the definition of trade union presence. No other variable on trade unions is available within the dataset and we had to rely on self-reported information and, since the definition of workplace union presence is self-reported, misclassification might happen. Furthermore, union presence does not guarantee *de facto* the presence of health and safety committees and translates into different workplaces’ practices ^47^. Finally, an important limitation is that we compared groups that are unlikely to be exchangeable with many potential residual confounding not measured in this study, as well as possible reverse causation.

This study takes place in a current climate of restrictions in the power of collective bargaining in countries where trade unions are present. Furthermore, despite a slight increase in union membership rates in the United Kingdom over the past two years, the long-term world trends show a massive decrease in union membership ^48^. Both individual behaviours and structural limitations have contributed to the current setting. The COVID-19 pandemic has illustrated the importance of work and employment in shaping population health and wellbeing. This study particularly demonstrates that, beyond individual unionization behaviours, collective negotiation at the workplace could have an alleviative mental health effect on the whole workforce and not only union members. This might encourage policy makers to promote collective bargaining and democracy at work to protect the workforce and improve population health by promoting individuals’ union membership on the one hand and facilitating collective negotiation on the other hand on the industrial / occupational level, especially during periods of crises.

## Data Availability

All data produced in the present study are available upon reasonable request to the authors
Original Understanding Society data are available upon request at: https://www.data-archive.ac.uk

## Supplementary files

S1. Flowchart of study participants -Transitions

S2. Main estimates (union presence and union membership – models 1, 2 and 3)

S3. Sample characteristics

S4. Sensitivity analyses:

Union presence – adjusted model (2-way interaction), excluding union members

Union presence – adjusted model (3-way interaction), excluding union members

Union presence – adjusted model (2-way interaction), excluding public workplaces

Union presence – adjusted model (3-way interaction), excluding public workplaces

Union presence – adjusted model (2-way interaction), GHQ-36

Union membership – adjusted model (2-way interaction), GHQ-36

## References

1 Cai M, Moore S, Ball C, Flynn M, Mulkearn K. The role of union health and safety representatives during the COVID-19 pandemic: A case study of the UK food processing, distribution, and retail sectors. Industrial Relations Journal 2022; 53: 390–407.

2 Wels J. The role of labour unions in explaining workers’ mental and physical health in Great Britain. A longitudinal approach. Soc Sci Med 2020; 247. DOI:10.1016/j.socscimed.2020.112796.

3 Eisenberg-Guyot J, Mooney SJ, Barrington WE, Hajat A. Does the Union Make Us Strong? Labor-Union Membership, Self-Rated Health, and Mental Illness: A Parametric G-Formula Approach. Am J Epidemiol 2020; 186: 227–36.

4. Wels J. RE: “Does the Union make us strong? Labor-Union membership, self-rated health, and mental illness: a parametric G-formula approach”. Am J Epidemiol 2021; 190: 1178–1178.

5 Sochas L, Reeves A. Does collective bargaining reduce health inequalities between labour market insiders and outsiders? Socioecon Rev 2022; published online Sept 1. DOI:10.1093/ser/mwac052.

6 Checchi D, Visser J. Pattern persistence in European trade union density: A longitudinal analysis 1950-1996. Eur Sociol Rev 2005; 21: 1–21.

7 Toubøl J, Jensen CS. Why do people join trade unions? The impact of workplace union density on union recruitment. Transfer: European Review of Labour and Research 2014; 20: 135–54.

8 Fazekas Z. Institutional effects on the presence of trade unions at the workplace: Moderation in a multilevel setting. European Journal of Industrial Relations 2011; 17: 153–69.

9 Department for Business Energy & Industrial Strategy. Trade Union Membership, UK 1995-2020: Statistical Bulletin. London: Crown, 2021.

10 Visser J. ICTWSS Data base. Version 6.1. Amsterdam: Amsterdam Institute for Advanced Labour Studies AIAS, 2019.

11 Wielgoszewska B, Booth C, Green MJ, Hamilton OK, Wels J. Association between home working and mental health by key worker status during the Covid-19 pandemic. Evidence from four British longitudinal studies. Ind Health 2022; 60: 1–12.

12 Wels J, Wielgoszewska B, Moltrecht B, et al. Home working and its association with social and mental wellbeing at different stages of the COVID-19 pandemic: Evidence from seven UK longitudinal population surveys. MedRxiv 2022; 2022.10.03.22280412: 1–27.

13 Wels J, Hamarat N. A shift in women’s health? Older workers’ self-reported health and employment settings during the COVID-19 pandemic. Eur J Public Health 2021; published online Nov 27. DOI:10.1093/eurpub/ckab204.

14 Wels J, Booth C, Wielgoszewska B, et al. Mental and social wellbeing and the UK coronavirus job retention scheme: Evidence from nine longitudinal studies. Soc Sci Med 2022; 308: 115226.

15 Patel K, Robertson E, Kwong ASF, et al. Psychological Distress Before and During the COVID-19 Pandemic Among Adults in the United Kingdom Based on Coordinated Analyses of 11 Longitudinal Studies. JAMA Netw Open 2022; 5: e227629.

16 Wels J, Wielgoszewska B, Moltrecht B, et al. Home working and social and mental wellbeing at different stages of the COVID-19 pandemic in the UK: Evidence from 7 longitudinal population surveys. PLoS Med 2023; 20: e1004214.

17. Mcnicholas C, Rhinehart L, Poydock M, Shierholz H, Perez D. Why unions are good for workers-especially in a crisis like COVID-19. 12 policies that would boost worker rights, safety, and wages. Economic Policy Institute 2020; 204014.

18 Hunt T, Connolly H. Covid-19 and the work of trade unions: Adaptation, transition and renewal. Industrial Relations Journal 2023; published online March 1. DOI:10.1111/irj.12395.

19 Watterson A. COVID-19 in the UK and Occupational Health and Safety: Predictable not Inevitable Failures by Government, and Trade Union and Nongovernmental Organization Responses. New Solutions 2020; 30: 86–94.

20 TUC. Covid-19: An Occupational Disease. 2022.

21 Brandl B. The cooperation between business organizations, trade unions, and the state during the COVID-19 pandemic: A comparative analysis of the nature of the tripartite relationship. Ind Relat (Berkeley) 2021; published online April 1. DOI:10.1111/irel.12300.

22 Bryson A. Health and safety risks in Britain’s workplaces: where are they and who controls them? Industrial Relations Journal 2016; 47: 547–66.

23 Han ES. What did unions do for union workers during the COVID-19 pandemic? Br J Ind Relat 2022. DOI:10.1111/bjir.12716.

24. Trade Union Congress. Workers’ experiences of long Covid A TUC report. 2021.

25 Wels J. Are there health benefits of being unionized in late career? A longitudinal approach using HRS. Am J Ind Med 2018; 61. DOI:10.1002/ajim.22877.

26 Niedzwiedz CL, Green MJ, Benzeval M, et al. Mental health and health behaviours before and during the initial phase of the COVID-19 lockdown: Longitudinal analyses of the UK Household Longitudinal Study. J Epidemiol Community Health (1978) 2021; 75: 224–31.

27 Kromydas T, Green M, Craig P, et al. Comparing population-level mental health of UK workers before and during the COVID-19 pandemic: A longitudinal study using Understanding Society. J Epidemiol Community Health (1978) 2022. DOI:10.1136/jech-2021-218561.

28 Wels J, Hamarat N. A shift in women’s health? Older workers’ self-reported health and employment settings during the COVID-19 pandemic. Eur J Public Health 2022; 32: 80–6.

29 Goldberg D. Manual of the general health questionnaire. Windsor: NFER Nelson, 1978.

30 Kelly MJ, Dunstan FD, Lloyd K, Fone DL. Evaluating cutpoints for the MHI-5 and MCS using the GHQ-12: a comparison of five different methods. BMC Psychiatry 2008; 8: 10.

31 Mutambudzi M, Niedwiedz C, Macdonald EB, et al. Occupation and risk of severe COVID-19: Prospective cohort study of 120 075 UK Biobank participants. Occup Environ Med 2021; 78: 307–14.

32. Ross CE, Mirowsky J. Does Employment Affect Health? 1995.

33 Reynolds MM, Buffel V. Organized Labor and Depression in Europe: Making Power Explicit in the Political Economy of Health. J Health Soc Behav 2020; 61: 342–58.

34 Taylor GS. A reanalysis of the relation between unionization and workplace safety. International Journal of Health Services 1987; 17: 443–53.

35 Appleton WC, Baker JG. The Effect of Unionization on Safety in Bituminous Deep Mines: Comment. J Labor Res 1985; 6: 209–10.

36 Kim WY, Cho HH. Unions, Health and Safety Committees, and Workplace Accidents in the Korean Manufacturing Sector. Saf Health Work 2016; 7: 161–5.

37 Wels J, Hamarat N, De Greef V. Social dialogue quality and workers’ health as perceived by Belgian trade union representatives during the COVID-19 pandemic. medRxiv 2023; 2023.04.10.23288317: 1–31.

38 Wels J. The role of labour unions in explaining workers’ mental and physical health in Great Britain. A longitudinal approach. Soc Sci Med 2020; 247: 112796.

39 Wels J. Are there health benefits of being unionized in late career? A longitudinal approach using HRS. Am J Ind Med 2018; 61: 751–61.

40 Freeman RB. Longitudinal Analyses of the Effects of Trade Unions. J Labor Econ 1984; 2: 1–26.

41 Bessa I, Charlwood A, Valizade D. Do Unions Cause Job Dissatisfaction? Evidence from a Quasi-Experiment. Br J Ind Relat 2020; 7: 1–29.

42 Topriceanu C-C, Wong A, Moon JC, et al. Impact of lockdown on key workers: findings from the COVID-19 survey in four UK national longitudinal studies. J Epidemiol Community Health (1978) 2021;: jech-2020-215889.

43 Adams-Prassl A, Boneva T, Golin M, Rauh C. Furloughing*. Fisc Stud 2020; 41: 591–622.

44 Hensher M. Covid-19, unemployment, and health: time for deeper solutions? BMJ 2020; 371: m3687.

45. Franklin P. ETUI Policy Brief. 2021.

46 Wels J. The Contribution of Labour Unions in Fostering Access to Flexible Work Arrangements in Britain. METICES Discussion Paper series 2021;: 22.

47 Moore S, Cai M, Ball C, Flynn M. Health and Safety Reps in COVID-19— Representation Unleashed? Int J Environ Res Public Health 2023; 20. DOI:10.3390/ijerph20085551.

48 Schnabel C. Union Membership and Collective Bargaining: Trends and Determinants. IZA discussion paper series 2020; 13465: 1–44.

