## Supplementary file S1-4 for "Trade unions and mental health during an employment crisis. Evidence from the UK before and during the COVID-19 pandemic"

**Supplementary files**

### Supplementary file 1. Flowchart of study participants

#### Union presence flowchart

Usoc observations (n) and participants (N) at baseline (Waves 9, 10/11 and all Covid-19 surveys:

|  | **Pre-Pandemic** | **Pandemic** | **Total** |
| --- | --- | --- | --- |
| **n** | 102,379 | 123,935 | **226,312** |
| **N** |  | | **41,287** |

n and N interviewed as proxies (Pre-pandemic)

|  | **Pre-Pandemic** | **Pandemic** | **Total** |
| --- | --- | --- | --- |
| **n** | -2,364 |  | **-2,364** |
| **N** |  | | **-1,145** |

Eligible n and N who were interviewed fully or partially:

|  | **Pre-Pandemic** | **Pandemic** | **Total** |
| --- | --- | --- | --- |
| **n** | 100,015 | 122,826 | **222,841** |
| **N** |  | | **40,142** |

n and N over 18y old:

|  | **Pre-Pandemic** | **Pandemic** | **Total** |
| --- | --- | --- | --- |
| **n** | -2,706 | -1,089 | **-3,795** |
| **N** |  | | **-914** |

Eligible n and N over 18y old:

|  | **Pre-Pandemic** | **Pandemic** | **Total** |
| --- | --- | --- | --- |
| **n** | 97,309 | 121,737 | **219,046** |
| **N** |  | | **39,228** |

n and N with missing or zero weight

|  | **Pre-Pandemic** | **Pandemic** | **Total** |
| --- | --- | --- | --- |
| **n** | -54,363 | -68,943 | **-123,306** |
| **N** |  | | **-11,867** |

Eligible n and N with non-missing cross-sectional weight

|  | **Pre-Pandemic** | **Pandemic** | **Total** |
| --- | --- | --- | --- |
| **n** | 42,946 | 52,794 | **95,740** |
| **N** |  | | **27,361** |

Eligible n and N with non-missing values in the outcome variable:

|  | **Pre-Pandemic** | **Pandemic** | **Total** |
| --- | --- | --- | --- |
| **n** | 42,607 | 51,269 | **93,876** |
| **N** |  | | **27,095** |

n and N with missing values in the outcome variable:

|  | **Pre-Pandemic** | **Pandemic** | **Total** |
| --- | --- | --- | --- |
| **n** | -339 | -1,525 | **-1,864** |
| **N** |  | | **-266** |

n and N with missing values in our key variables who are self-employed and not in employment :

|  | **Pre-Pandemic** | **Pandemic** | **Total** |
| --- | --- | --- | --- |
| **n** | -16,000 | -5,545 | **-21,545** |
| **N** |  | | **-13,004** |

Eligible n and N in dependent employment with no missing values in key variables*:

|  | **Pre-Pandemic** | **Pandemic** | **Total** |
| --- | --- | --- | --- |
| **n** | 26,946 | 47,249 | **74,195** |
| **N** |  | | **14,091** |

*: Outcome, exposures and all covariates used in Models 1 and 2

Eligible n and N who participated in wave 9 and at least one Covid-19 survey:

|  | **Pre-Pandemic** | **Pandemic** | **Total** |
| --- | --- | --- | --- |
| **n** | 14,142 | 35,773 | **49,915** |
| **N** |  | | **5,988** |

n and N that only participated in either pandemic or Covid-19 surveys:

|  | **Pre-Pandemic** | **Pandemic** | **Total** |
| --- | --- | --- | --- |
| **n** | -28,465 | -15,496 | **-43,961** |
| **N** |  | | **-8,103** |

#### Union membership flowchart

Usoc observations (n) and participants (N) at baseline (Waves 9, 10/11 and all Covid-19 surveys:

|  | **Pre-Pandemic** | **Pandemic** | **Total** |
| --- | --- | --- | --- |
| **n** | 102,379 | 123,935 | **226,314** |
| **N** |  | | **41,287** |

n and N interviewed as proxies (Pre-pandemic)

|  | **Pre-Pandemic** | **Pandemic** | **Total** |
| --- | --- | --- | --- |
| **n** | -2,364 |  | **-2,364** |
| **N** |  | | **-1,145** |

Eligible n and N who were interviewed fully or partially:

|  | **Pre-Pandemic** | **Pandemic** | **Total** |
| --- | --- | --- | --- |
| **n** | 100,015 | 122,826 | **222,841** |
| **N** |  | | **40,142** |

n and N over 18y old:

|  | **Pre-Pandemic** | **Pandemic** | **Total** |
| --- | --- | --- | --- |
| **n** | -2,706 | -1,089 | **-3,795** |
| **N** |  | | **-914** |

Eligible n and N over 18y old:

|  | **Pre-Pandemic** | **Pandemic** | **Total** |
| --- | --- | --- | --- |
| **n** | 97,309 | 121,737 | **219,046** |
| **N** |  | | **39,228** |

n and N with missing or zero weight

|  | **Pre-Pandemic** | **Pandemic** | **Total** |
| --- | --- | --- | --- |
| **n** | -83,200 | -92,625 | **-175,825** |
| **N** |  | | **-31,698** |

Eligible n and N with non-missing cross-sectional weight

|  | **Pre-Pandemic** | **Pandemic** | **Total** |
| --- | --- | --- | --- |
| **n** | 14,109 | 29,112 | **43,221** |
| **N** |  | | **27,361** |

Eligible n and N with non-missing values in the outcome variable:

|  | **Pre-Pandemic** | **Pandemic** | **Total** |
| --- | --- | --- | --- |
| **n** | 14,024 | 28,306 | **42,330** |
| **N** |  | | **7,466** |

n and N with missing values in the outcome variable:

|  | **Pre-Pandemic** | **Pandemic** | **Total** |
| --- | --- | --- | --- |
| **n** | -85 | -806 | **-891** |
| **N** |  | | **-64** |

n and N with missing values in our key variables who are self-employed and not in employment :

|  | **Pre-Pandemic** | **Pandemic** | **Total** |
| --- | --- | --- | --- |
| **n** | -525 | -1812 | **-2,337** |
| **N** |  | | **-230** |

Eligible n and N in dependent employment with no missing values in key variables*:

|  | **Pre-Pandemic** | **Pandemic** | **Total** |
| --- | --- | --- | --- |
| **n** | 13,499 | 26,494 | **39,993** |
| **N** |  | | **7,236** |

*: Outcome, exposures and all covariates used in Models 1 and 2

n and N that only participated in either pandemic or Covid-19 surveys:

|  | **Pre-Pandemic** | **Pandemic** | **Total** |
| --- | --- | --- | --- |
| **n** | -6,032 | -5,990 | **-12,022** |
| **N** |  | | **-3,895** |

Figure 2: Flowchart for union membership

Eligible n and N who participated in wave 9 and at least one Covid-19 survey:

|  | **Pre-Pandemic** | **Pandemic** | **Total** |
| --- | --- | --- | --- |
| **n** | 7,467 | 20,504 | **27,971** |
| **N** |  | | **3,341** |

#### Testing the stability assumption (Trade Union presence and Trade union membership variables)

Trade union presence and membership variables are asked every two waves and therefore data exist only in waves 2,4,6,8 and 10. Using a sample that includes only these waves we first calculated a variable that shows the change in non-missing values of the trade union presence and membership variables between two consecutive waves. Then we cross-tabulated it over the original variable, again excluding all its missing values. We found that respondents in unionised workplaces and non-unionised workplaces have a very high and also very similar overall stability of 87% and 86%, respectively. For trade union members the equivalent percentages are 93% and 84% for trade union members and non-members, respectively (Tables S3 and S4).

##### Table S1:Transitions between trade union presence values (Yes :Unionised workplace, No: Non-unionised workplace)

|  | Frequency | Percentage |
| --- | --- | --- |
| Stable between two waves (non-unionised workplaces) - From No to No | 21,961 | 85.81 |
| Transition from a Unionised to a non-unionised workplace – From Yes to No | 3,632 | 14.19 |
| **Total (non-unionised workplaces)** | **25,593** | **100.00** |
| Stable between two waves (unionised workplaces) - From Yes to Yes | 23,408 | 86.8 |
| Transition from a non-Unionised to a unionised workplace – From No to Yes | 3,561 | 13.20 |
| **Total (unionised workplaces)** | **26,969** | **100.00** |

##### Table S2:Transitions between trade union membership values (Yes :member, No: Non-member)

|  | Frequency | Percentage |
| --- | --- | --- |
| Stable between two waves (non-member) - From No to No | 7,104 | 84.36 |
| Transition from member to a non-member – From Yes to No | 1,317 | 15.64 |
| **Total (non-member )** | **8,421** | **100.00** |
| Stable between two waves (member) - From Yes to Yes | 13,884 | 92.94 |
| Transition from a non-member to member – From No to Yes | 1,054 | 7.06 |
| **Total (member)** | **14,938** | **100.00** |

### Supplementary file S2 – Main estimates

#### Union presence – unadjusted model

|  |  | **Unadjusted model (weighted)** | | | | | |
| --- | --- | --- | --- | --- | --- | --- | --- |
|  |  | **OR** | **std. err.** | **z** | **P>z** | **[95% conf.** | **interval]** |
| Pre-Post Pandemic | | 1.31 | 0.14 | 2.47 | 0.01 | 1.06 | 1.62 |
| Union presence: yes | | 1.21 | 0.10 | 2.27 | 0.02 | 1.03 | 1.43 |
| Pre-Post Pandemic # union presence: yes | | 0.86 | 0.08 | -1.61 | 0.11 | 0.71 | 1.03 |
| Wave Number: | |  |  |  |  |  |  |
|  | Wave 10 | 0.90 | 0.07 | -1.36 | 0.17 | 0.77 | 1.05 |
|  | Wave 11 | 1.19 | 0.10 | 2.07 | 0.04 | 1.01 | 1.41 |
|  | Apr-20 | 2.34 | 0.21 | 9.46 | 0.00 | 1.96 | 2.79 |
|  | **COVID-19 sweeps** |  |  |  |  |  |  |
|  | May-20 | 1.85 | 0.17 | 6.76 | 0.00 | 1.55 | 2.21 |
|  | Jun-20 | 1.63 | 0.16 | 5.00 | 0.00 | 1.34 | 1.97 |
|  | Jul-20 | 0.99 | 0.09 | -0.14 | 0.89 | 0.82 | 1.18 |
|  | Sep-20 | 1.06 | 0.10 | 0.62 | 0.53 | 0.88 | 1.29 |
|  | Nov-20 | 1.85 | 0.19 | 6.11 | 0.00 | 1.52 | 2.25 |
|  | Jan-21 | 1.81 | 0.17 | 6.24 | 0.00 | 1.50 | 2.18 |
|  | Mar-21 | 1.25 | 0.12 | 2.30 | 0.02 | 1.03 | 1.50 |
|  | Sep-21 | 1.00 | (omitted) |  |  |  |  |
| Constant | | 0.08 | 0.01 | -30.08 | 0.00 | 0.06 | 0.09 |

Note: Observations=49,915; Groups=5,988 (Minimum=1; Average= 8.3; Maximum=12).

#### Union presence – adjusted model (2-way interaction)

|  |  | **Adjusted model (weighted)** | | | | | |
| --- | --- | --- | --- | --- | --- | --- | --- |
|  |  | **OR** | **std. err.** | **z** | **P>z** | **[95% conf.** | **interval]** |
| Sex |  | 2.16 | 0.15 | 11.42 | 0.00 | 1.89 | 2.47 |
| Age |  | 1.01 | 0.02 | 0.32 | 0.75 | 0.97 | 1.04 |
| Age_square | | 1.00 | 0.00 | -1.57 | 0.12 | 1.00 | 1.00 |
| Ethnicity | | 0.90 | 0.11 | -0.85 | 0.40 | 0.72 | 1.14 |
| Pre-Post Pandemic | | 1.45 | 0.16 | 3.34 | 0.00 | 1.17 | 1.80 |
| Union presence: yes | | 1.11 | 0.10 | 1.18 | 0.24 | 0.93 | 1.33 |
| Pre-Post Pandemic # Union presence: yes | | 0.89 | 0.09 | -1.23 | 0.22 | 0.73 | 1.07 |
| Industry: | |  |  |  |  |  |  |
|  | Mining, Energy and Water Supply | 1.13 | 0.29 | 0.47 | 0.64 | 0.68 | 1.86 |
|  | Manufacturing | 0.66 | 0.13 | -2.16 | 0.03 | 0.45 | 0.96 |
|  | Construction | 0.68 | 0.15 | -1.75 | 0.08 | 0.44 | 1.05 |
|  | Wholesale and Retail Trade Motor Repair | 0.85 | 0.16 | -0.90 | 0.37 | 0.59 | 1.22 |
|  | Transportation and Storage | 0.68 | 0.15 | -1.80 | 0.07 | 0.44 | 1.04 |
|  | Information and Communication | 0.85 | 0.18 | -0.77 | 0.44 | 0.57 | 1.28 |
|  | Financial and Insurance Activities | 0.64 | 0.14 | -2.03 | 0.04 | 0.42 | 0.98 |
|  | Real Estate Activities | 0.96 | 0.27 | -0.16 | 0.88 | 0.55 | 1.65 |
|  | Professional Scientific and Technical | 0.79 | 0.15 | -1.24 | 0.21 | 0.55 | 1.14 |
|  | Administrative and Support Services | 0.62 | 0.13 | -2.32 | 0.02 | 0.41 | 0.93 |
|  | Public Administration and Defence | 0.75 | 0.14 | -1.54 | 0.12 | 0.52 | 1.08 |
|  | Education | 0.78 | 0.14 | -1.36 | 0.17 | 0.55 | 1.11 |
|  | Human Health and Social Work Activities | 0.81 | 0.14 | -1.21 | 0.23 | 0.57 | 1.14 |
|  | Other Services | 0.67 | 0.13 | -2.12 | 0.03 | 0.47 | 0.97 |
| Country: | |  |  |  |  |  |  |
|  | Wales | 1.24 | 0.16 | 1.70 | 0.09 | 0.97 | 1.59 |
|  | Scotland | 0.98 | 0.11 | -0.17 | 0.86 | 0.79 | 1.22 |
|  | Northern Ireland | 0.87 | 0.13 | -0.92 | 0.36 | 0.65 | 1.17 |
| Highest level of education: | |  |  |  |  |  |  |
|  | A-Level | 0.73 | 0.06 | -4.01 | 0.00 | 0.62 | 0.85 |
|  | GCSE | 0.75 | 0.07 | -3.20 | 0.00 | 0.63 | 0.90 |
|  | Other | 0.55 | 0.09 | -3.50 | 0.00 | 0.39 | 0.77 |
|  | None | 0.37 | 0.12 | -3.19 | 0.00 | 0.20 | 0.68 |
| Self-reported financial conditions: bad | | 2.44 | 0.16 | 13.45 | 0.00 | 2.14 | 2.78 |
| Health condition: yes | | 2.09 | 0.14 | 11.25 | 0.00 | 1.84 | 2.38 |
| Company size: | |  |  |  |  |  |  |
|  | 25-199 | 0.89 | 0.07 | -1.46 | 0.14 | 0.76 | 1.04 |
|  | More than 200 | 0.95 | 0.07 | -0.64 | 0.52 | 0.81 | 1.11 |
| Wave Number | |  |  |  |  |  |  |
|  | Wave 10 | 0.91 | 0.07 | -1.23 | 0.22 | 0.78 | 1.06 |
|  | Wave 11 | 1.22 | 0.11 | 2.32 | 0.02 | 1.03 | 1.45 |
|  | **COVID-19 sweeps** |  |  |  |  |  |  |
|  | Apr-20 | 2.29 | 0.20 | 9.22 | 0.00 | 1.92 | 2.73 |
|  | May-20 | 1.81 | 0.17 | 6.47 | 0.00 | 1.51 | 2.16 |
|  | Jun-20 | 1.62 | 0.16 | 4.98 | 0.00 | 1.34 | 1.96 |
|  | Jul-20 | 0.96 | 0.09 | -0.44 | 0.66 | 0.80 | 1.15 |
|  | Sep-20 | 1.04 | 0.10 | 0.42 | 0.68 | 0.86 | 1.26 |
|  | Nov-20 | 1.81 | 0.18 | 5.82 | 0.00 | 1.48 | 2.21 |
|  | Jan-21 | 1.78 | 0.17 | 5.99 | 0.00 | 1.47 | 2.15 |
|  | Mar-21 | 1.27 | 0.12 | 2.43 | 0.02 | 1.05 | 1.54 |
|  | Sep-21 | 1.00 | (omitted) |  |  |  |  |
| Constant | | 0.04 | 0.02 | -7.06 | 0.00 | 0.02 | 0.10 |

Note: Observations=49,915; Groups=5,988 (Minimum=1; Average= 8.3; Maximum=12).

#### Union presence – adjusted model (3-way interaction)

|  |  | **Adjusted model (weighted) -- three-ways interaction** | | | | | |
| --- | --- | --- | --- | --- | --- | --- | --- |
|  |  | **OR** | **std. err.** | **z** | **P>z** | **[95% conf.** | **interval]** |
| Sex |  | 2.15 | 0.15 | 11.27 | 0.00 | 1.88 | 2.45 |
| Age |  | 1.01 | 0.02 | 0.32 | 0.75 | 0.97 | 1.04 |
| Age square | | 1.00 | 0.00 | -1.59 | 0.11 | 1.00 | 1.00 |
| Ethniciy | | 0.90 | 0.11 | -0.90 | 0.37 | 0.71 | 1.13 |
| Pre-Post Pandemic # Industry: |  |  |  |  |  |  |  |
|  | Pre-Pandemic-#Mining, Energy and Water Supply | 2.81 | 1.57 | 1.85 | 0.07 | 0.94 | 8.41 |
|  | Post-Pandemic-#Mining, Energy and Water Supply | 2.07 | 0.64 | 2.37 | 0.02 | 1.13 | 3.79 |
|  | Pre-Pandemic-#Manufacturing | 0.82 | 0.20 | -0.81 | 0.42 | 0.51 | 1.32 |
|  | Post-Pandemic-#Manufacturing | 1.29 | 0.31 | 1.06 | 0.29 | 0.81 | 2.05 |
|  | Pre-Pandemic-#Construction | 0.67 | 0.21 | -1.25 | 0.21 | 0.36 | 1.25 |
|  | Post-Pandemic-#Construction | 1.11 | 0.29 | 0.41 | 0.68 | 0.67 | 1.86 |
|  | Pre-Pandemic-#Wholesale and Retail Trade Motor Repair | 1.04 | 0.26 | 0.14 | 0.89 | 0.64 | 1.69 |
|  | Post-Pandemic-#Wholesale and Retail Trade Motor Repair | 1.53 | 0.37 | 1.75 | 0.08 | 0.95 | 2.46 |
|  | Pre-Pandemic-#Transportation and Storage | 0.92 | 0.34 | -0.23 | 0.82 | 0.45 | 1.88 |
|  | Post-Pandemic-#Transportation and Storage | 0.85 | 0.28 | -0.49 | 0.63 | 0.44 | 1.63 |
|  | Pre-Pandemic-#Accommodation and Food Services | 1.59 | 0.49 | 1.51 | 0.13 | 0.87 | 2.91 |
|  | Post-Pandemic-#Accommodation and Food Services | 1.65 | 0.48 | 1.71 | 0.09 | 0.93 | 2.92 |
|  | Pre-Pandemic-#Information and Communication | 0.71 | 0.25 | -0.96 | 0.34 | 0.36 | 1.42 |
|  | Post-Pandemic-#Information and Communication | 1.59 | 0.37 | 2.01 | 0.05 | 1.01 | 2.50 |
|  | Pre-Pandemic-#Financial and Insurance Activities | 1.43 | 0.43 | 1.20 | 0.23 | 0.80 | 2.56 |
|  | Post-Pandemic-#Financial and Insurance Activities | 1.03 | 0.26 | 0.12 | 0.90 | 0.63 | 1.70 |
|  | Pre-Pandemic-#Real Estate Activities | 0.81 | 0.44 | -0.39 | 0.70 | 0.28 | 2.33 |
|  | Post-Pandemic-#Real Estate Activities | 1.54 | 0.61 | 1.10 | 0.27 | 0.71 | 3.35 |
|  | Pre-Pandemic-#Professional Scientific and Technical | 0.83 | 0.19 | -0.79 | 0.43 | 0.53 | 1.30 |
|  | Post-Pandemic-#Professional Scientific and Technical | 1.37 | 0.30 | 1.44 | 0.15 | 0.89 | 2.09 |
|  | Pre-Pandemic-#Administrative and Support Services | 0.58 | 0.20 | -1.59 | 0.11 | 0.29 | 1.14 |
|  | Post-Pandemic-#Administrative and Support Services | 1.05 | 0.28 | 0.19 | 0.85 | 0.63 | 1.76 |
|  | Pre-Pandemic-#Public Administration and Defence | 0.43 | 0.26 | -1.38 | 0.17 | 0.13 | 1.42 |
|  | Post-Pandemic-#Public Administration and Defence | 1.07 | 0.33 | 0.21 | 0.83 | 0.58 | 1.96 |
|  | Pre-Pandemic-#Education | 0.51 | 0.17 | -2.03 | 0.04 | 0.27 | 0.98 |
|  | Post-Pandemic-#Education | 1.47 | 0.35 | 1.61 | 0.11 | 0.92 | 2.34 |
|  | Post-Pandemic-#Human Health and Social Work Activities | 1.22 | 0.25 | 0.99 | 0.32 | 0.82 | 1.81 |
|  | Pre-Pandemic-#Other Services | 1.15 | 0.36 | 0.45 | 0.65 | 0.62 | 2.14 |
|  | Post-Pandemic-#Other Services | 1.18 | 0.25 | 0.79 | 0.43 | 0.78 | 1.79 |
| Union presence: yes | | 1.10 | 0.21 | 0.51 | 0.61 | 0.76 | 1.60 |
| Pre-Post Pandemic # Industry # Union presence | |  |  |  |  |  |  |
|  | Pre-Pandemic-#Mining, Energy and Water Supply#Yes | 0.48 | 0.35 | -1.02 | 0.31 | 0.11 | 1.98 |
|  | Post-Pandemic-#Mining, Energy and Water Supply#Yes | 0.57 | 0.28 | -1.15 | 0.25 | 0.22 | 1.48 |
|  | Pre-Pandemic-#Manufacturing#Yes | 0.60 | 0.23 | -1.33 | 0.18 | 0.28 | 1.28 |
|  | Post-Pandemic-#Manufacturing#Yes | 0.56 | 0.20 | -1.66 | 0.10 | 0.28 | 1.11 |
|  | Pre-Pandemic-#Construction#Yes | 2.09 | 1.16 | 1.32 | 0.19 | 0.70 | 6.22 |
|  | Post-Pandemic-#Construction#Yes | 0.99 | 0.41 | -0.02 | 0.99 | 0.45 | 2.22 |
|  | Pre-Pandemic-#Wholesale and Retail Trade Motor Repair#Yes | 0.98 | 0.33 | -0.06 | 0.95 | 0.51 | 1.90 |
|  | Post-Pandemic-#Wholesale and Retail Trade Motor Repair#Yes | 0.71 | 0.21 | -1.14 | 0.25 | 0.39 | 1.28 |
|  | Pre-Pandemic-#Transportation and Storage#Yes | 0.76 | 0.34 | -0.61 | 0.54 | 0.32 | 1.82 |
|  | Post-Pandemic-#Transportation and Storage#Yes | 1.48 | 0.59 | 0.99 | 0.32 | 0.68 | 3.25 |
|  | Pre-Pandemic-#Accommodation and Food Services#Yes | 0.30 | 0.19 | -1.91 | 0.06 | 0.09 | 1.03 |
|  | Post-Pandemic-#Accommodation and Food Services#Yes | 0.89 | 0.39 | -0.27 | 0.79 | 0.38 | 2.10 |
|  | Pre-Pandemic-#Information and Communication#Yes | 3.08 | 1.85 | 1.86 | 0.06 | 0.94 | 10.02 |
|  | Post-Pandemic-#Information and Communication#Yes | 0.72 | 0.25 | -0.94 | 0.35 | 0.36 | 1.43 |
|  | Pre-Pandemic-#Financial and Insurance Activities#Yes | 0.54 | 0.24 | -1.38 | 0.17 | 0.22 | 1.30 |
|  | Post-Pandemic-#Financial and Insurance Activities#Yes | 0.74 | 0.26 | -0.86 | 0.39 | 0.37 | 1.47 |
|  | Pre-Pandemic-#Real Estate Activities#Yes | 4.32 | 3.03 | 2.08 | 0.04 | 1.09 | 17.09 |
|  | Post-Pandemic-#Real Estate Activities#Yes | 0.48 | 0.31 | -1.15 | 0.25 | 0.13 | 1.68 |
|  | Pre-Pandemic-#Professional Scientific and Technical#Yes | 0.95 | 0.36 | -0.14 | 0.89 | 0.45 | 2.01 |
|  | Post-Pandemic-#Professional Scientific and Technical#Yes | 1.00 | 0.27 | -0.01 | 0.99 | 0.59 | 1.69 |
|  | Pre-Pandemic-#Administrative and Support Services#Yes | 1.12 | 0.69 | 0.18 | 0.86 | 0.33 | 3.74 |
|  | Post-Pandemic-#Administrative and Support Services#Yes | 0.99 | 0.32 | -0.03 | 0.97 | 0.52 | 1.88 |
|  | Pre-Pandemic-#Public Administration and Defence#Yes | 2.11 | 1.33 | 1.19 | 0.23 | 0.62 | 7.23 |
|  | Post-Pandemic-#Public Administration and Defence#Yes | 1.16 | 0.39 | 0.45 | 0.66 | 0.60 | 2.23 |
|  | Pre-Pandemic-#Education#Yes | 1.74 | 0.62 | 1.56 | 0.12 | 0.87 | 3.51 |
|  | Post-Pandemic-#Education#Yes | 0.86 | 0.22 | -0.58 | 0.56 | 0.52 | 1.43 |
|  | Post-Pandemic-#Human Health and Social Work Activities#Yes | 1.09 | 0.25 | 0.39 | 0.69 | 0.70 | 1.70 |
|  | Pre-Pandemic-#Other Services #Yes | 0.67 | 0.31 | -0.85 | 0.40 | 0.27 | 1.68 |
|  | Post-Pandemic-#Other Services #Yes | 0.80 | 0.22 | -0.81 | 0.42 | 0.47 | 1.36 |
| Country: | |  |  |  |  |  |  |
|  | Wales | 1.25 | 0.16 | 1.76 | 0.08 | 0.97 | 1.61 |
|  | Scotland | 0.98 | 0.11 | -0.22 | 0.83 | 0.78 | 1.22 |
|  | Northern Ireland | 0.87 | 0.13 | -0.94 | 0.35 | 0.65 | 1.16 |
| Highest level of education: | |  |  |  |  |  |  |
|  | A-Level | 0.74 | 0.06 | -3.81 | 0.00 | 0.63 | 0.86 |
|  | GCSE | 0.77 | 0.07 | -2.97 | 0.00 | 0.64 | 0.91 |
|  | Other | 0.55 | 0.10 | -3.45 | 0.00 | 0.39 | 0.77 |
|  | None | 0.38 | 0.12 | -3.07 | 0.00 | 0.20 | 0.70 |
| Self-reported financial conditions: bad | | 2.45 | 0.16 | 13.45 | 0.00 | 2.15 | 2.79 |
| Health condition: yes | | 2.08 | 0.14 | 11.18 | 0.00 | 1.83 | 2.37 |
| Company size: | |  |  |  |  |  |  |
|  | 25-199 | 0.90 | 0.07 | -1.38 | 0.17 | 0.77 | 1.05 |
|  | More than 200 | 0.96 | 0.08 | -0.55 | 0.59 | 0.82 | 1.12 |
| Wave Number | |  |  |  |  |  |  |
|  | Wave 10 | 0.91 | 0.07 | -1.26 | 0.21 | 0.78 | 1.06 |
|  | Wave 11 | 1.24 | 0.11 | 2.51 | 0.01 | 1.05 | 1.47 |
|  | **COVID-19 sweeps** |  |  |  |  |  |  |
|  | Apr-20 | 2.29 | 0.21 | 9.25 | 0.00 | 1.92 | 2.73 |
|  | May-20 | 1.81 | 0.17 | 6.50 | 0.00 | 1.51 | 2.17 |
|  | Jun-20 | 1.62 | 0.16 | 5.00 | 0.00 | 1.34 | 1.96 |
|  | Jul-20 | 0.96 | 0.09 | -0.46 | 0.64 | 0.80 | 1.15 |
|  | Sep-20 | 1.04 | 0.10 | 0.38 | 0.71 | 0.86 | 1.26 |
|  | Nov-20 | 1.81 | 0.18 | 5.81 | 0.00 | 1.48 | 2.21 |
|  | Jan-21 | 1.77 | 0.17 | 5.95 | 0.00 | 1.47 | 2.14 |
|  | Mar-21 | 1.27 | 0.12 | 2.44 | 0.02 | 1.05 | 1.54 |
|  | Sep-21 | 1.00 | (omitted) |  |  |  |  |
| Constant | | 0.04 | 0.02 | -7.22 | 0.00 | 0.01 | 0.09 |

Note: Observations=49,915; Groups=5,988 (Minimum=1; Average= 8.3; Maximum=12).

#### Union membership – unadjusted model

|  |  | **Unadjusted model (weighted)** | | | | | |
| --- | --- | --- | --- | --- | --- | --- | --- |
|  |  | **OR** | **std. err.** | **z** | **P>z** | **[95% conf.** | **interval]** |
| Pre-Post Pandemic | | 1.24 | 0.17 | 1.57 | 0.12 | 0.95 | 1.63 |
| Union membership: yes | | 1.24 | 0.15 | 1.85 | 0.06 | 0.99 | 1.56 |
| Pre-Post Pandemic # union membership: yes | | 0.85 | 0.11 | -1.27 | 0.21 | 0.66 | 1.09 |
| Wave Number: | |  |  |  |  |  |  |
|  | Wave 10 | 0.88 | 0.09 | -1.24 | 0.22 | 0.71 | 1.08 |
|  | Wave 11 | 1.32 | 0.16 | 2.35 | 0.02 | 1.05 | 1.67 |
|  | **COVID-19 sweeps** |  |  |  |  |  |  |
|  | Apr-20 | 2.54 | 0.28 | 8.44 | 0.00 | 2.05 | 3.15 |
|  | May-20 | 1.77 | 0.20 | 4.94 | 0.00 | 1.41 | 2.22 |
|  | Jun-20 | 1.49 | 0.18 | 3.25 | 0.00 | 1.17 | 1.90 |
|  | Jul-20 | 0.87 | 0.10 | -1.18 | 0.24 | 0.69 | 1.10 |
|  | Sep-20 | 1.12 | 0.13 | 0.92 | 0.36 | 0.88 | 1.41 |
|  | Nov-20 | 1.97 | 0.24 | 5.53 | 0.00 | 1.55 | 2.50 |
|  | Jan-21 | 2.22 | 0.25 | 7.05 | 0.00 | 1.78 | 2.77 |
|  | Mar-21 | 1.24 | 0.15 | 1.75 | 0.08 | 0.97 | 1.58 |
|  | Sep-21 | 1.00 | (omitted) |  |  |  |  |
| Constant | | 0.08 | 0.01 | -21.86 | 0.00 | 0.07 | 0.10 |

Note: Observations=27,971; Groups=3,341 (Minimum=1; Average= 8.4; Maximum=12).

#### Union membership – adjusted model (2-ways interaction)

|  |  | **Adjusted model (weighted)** | | | | | |
| --- | --- | --- | --- | --- | --- | --- | --- |
|  |  | **OR** | **std. err.** | **z** | **P>z** | **[95% conf.** | **interval]** |
| Sex |  | 2.08 | 0.20 | 7.81 | 0.00 | 1.73 | 2.50 |
| Age |  | 1.02 | 0.03 | 0.77 | 0.44 | 0.97 | 1.08 |
| Age_square | | 1.00 | 0.00 | -1.32 | 0.19 | 1.00 | 1.00 |
| Ethnicity | | 0.82 | 0.12 | -1.40 | 0.16 | 0.61 | 1.08 |
| Pre-Post Pandemic | | 1.40 | 0.19 | 2.43 | 0.02 | 1.07 | 1.83 |
| Union membership: yes | | 1.17 | 0.14 | 1.37 | 0.17 | 0.93 | 1.47 |
| Pre-Post Pandemic # Union membership: yes | | 0.84 | 0.11 | -1.35 | 0.18 | 0.66 | 1.08 |
| Industry: | |  |  |  |  |  |  |
|  | Mining, Energy and Water Supply | 1.08 | 0.44 | 0.20 | 0.84 | 0.49 | 2.42 |
|  | Manufacturing | 0.62 | 0.20 | -1.46 | 0.14 | 0.33 | 1.18 |
|  | Construction | 1.08 | 0.42 | 0.21 | 0.84 | 0.51 | 2.32 |
|  | Wholesale and Retail Trade Motor Repair | 1.07 | 0.32 | 0.21 | 0.83 | 0.59 | 1.93 |
|  | Transportation and Storage | 1.02 | 0.31 | 0.08 | 0.94 | 0.56 | 1.86 |
|  | Information and Communication | 1.18 | 0.40 | 0.49 | 0.63 | 0.61 | 2.30 |
|  | Financial and Insurance Activities | 0.63 | 0.21 | -1.37 | 0.17 | 0.32 | 1.22 |
|  | Real Estate Activities | 1.35 | 0.51 | 0.80 | 0.43 | 0.65 | 2.82 |
|  | Professional Scientific and Technical | 1.16 | 0.34 | 0.51 | 0.61 | 0.65 | 2.06 |
|  | Administrative and Support Services | 0.82 | 0.26 | -0.63 | 0.53 | 0.44 | 1.52 |
|  | Public Administration and Defence | 1.00 | 0.27 | 0.00 | 1.00 | 0.58 | 1.71 |
|  | Education | 1.05 | 0.28 | 0.19 | 0.85 | 0.62 | 1.77 |
|  | Human Health and Social Work Activities | 1.11 | 0.29 | 0.40 | 0.69 | 0.66 | 1.87 |
|  | Other Services | 0.80 | 0.23 | -0.78 | 0.44 | 0.45 | 1.41 |
| Country: | |  |  |  |  |  |  |
|  | Wales | 1.04 | 0.17 | 0.26 | 0.79 | 0.76 | 1.43 |
|  | Scotland | 0.94 | 0.12 | -0.49 | 0.62 | 0.73 | 1.21 |
|  | Northern Ireland | 0.89 | 0.16 | -0.66 | 0.51 | 0.63 | 1.26 |
| Highest level of education: | |  |  |  |  |  |  |
|  | A-Level | 0.75 | 0.08 | -2.66 | 0.01 | 0.61 | 0.93 |
|  | GCSE | 0.64 | 0.08 | -3.58 | 0.00 | 0.50 | 0.82 |
|  | Other | 0.45 | 0.11 | -3.34 | 0.00 | 0.28 | 0.72 |
|  | None | 0.42 | 0.18 | -2.00 | 0.05 | 0.18 | 0.98 |
| Self-reported financial conditions: bad | | 2.74 | 0.22 | 12.30 | 0.00 | 2.33 | 3.22 |
| Health condition: yes | | 1.94 | 0.17 | 7.57 | 0.00 | 1.63 | 2.30 |
| Company size: | |  |  |  |  |  |  |
|  | 25-199 | 0.95 | 0.11 | -0.45 | 0.65 | 0.75 | 1.19 |
|  | More than 200 | 1.05 | 0.12 | 0.48 | 0.63 | 0.85 | 1.31 |
| Wave Number | |  |  |  |  |  |  |
|  | Wave 10 | 0.89 | 0.09 | -1.12 | 0.26 | 0.72 | 1.09 |
|  | Wave 11 | 1.35 | 0.16 | 2.51 | 0.01 | 1.07 | 1.70 |
|  | **COVID-19 sweeps** |  |  |  |  |  |  |
|  | Apr-20 | 2.57 | 0.28 | 8.63 | 0.00 | 2.07 | 3.18 |
|  | May-20 | 1.78 | 0.20 | 5.02 | 0.00 | 1.42 | 2.23 |
|  | Jun-20 | 1.55 | 0.19 | 3.55 | 0.00 | 1.22 | 1.98 |
|  | Jul-20 | 0.87 | 0.10 | -1.19 | 0.23 | 0.69 | 1.10 |
|  | Sep-20 | 1.13 | 0.13 | 0.99 | 0.32 | 0.89 | 1.42 |
|  | Nov-20 | 1.95 | 0.24 | 5.35 | 0.00 | 1.53 | 2.50 |
|  | Jan-21 | 2.23 | 0.25 | 7.02 | 0.00 | 1.78 | 2.79 |
|  | Mar-21 | 1.27 | 0.16 | 1.93 | 0.05 | 1.00 | 1.62 |
|  | Sep-21 | 1.00 | (omitted) |  |  |  |  |
| Constant | | 0.02 | 0.01 | -5.65 | 0.00 | 0.01 | 0.08 |

Note: Observations=27,971; Groups=3,341 (Minimum=1; Average= 8.4; Maximum=12).

#### Union membership – adjusted model (3-ways interaction)

|  |  | **Adjusted model (weighted) -- three-ways interaction** | | | | | |
| --- | --- | --- | --- | --- | --- | --- | --- |
|  |  | **OR** | **std. err.** | **z** | **P>z** | **[95% conf.** | **interval]** |
| Sex |  | 2.11 | 0.20 | 7.90 | 0.00 | 1.75 | 2.54 |
| Age |  | 1.02 | 0.03 | 0.74 | 0.46 | 0.97 | 1.08 |
| Age square | | 1.00 | 0.00 | -1.29 | 0.20 | 1.00 | 1.00 |
| Ethniciy | | 0.81 | 0.12 | -1.44 | 0.15 | 0.61 | 1.08 |
| Pre-Post Pandemic # Industry: |  |  |  |  |  |  |  |
|  | Pre-Pandemic-#Mining, Energy and Water Supply | 0.98 | 0.79 | -0.02 | 0.98 | 0.20 | 4.77 |
|  | Post-Pandemic-#Mining, Energy and Water Supply | 1.13 | 0.69 | 0.19 | 0.85 | 0.34 | 3.77 |
|  | Pre-Pandemic-#Manufacturing | 0.25 | 0.15 | -2.33 | 0.02 | 0.08 | 0.80 |
|  | Post-Pandemic-#Manufacturing | 0.64 | 0.26 | -1.08 | 0.28 | 0.29 | 1.44 |
|  | Pre-Pandemic-#Construction | 1.79 | 1.09 | 0.96 | 0.34 | 0.54 | 5.91 |
|  | Post-Pandemic-#Construction | 0.88 | 0.39 | -0.30 | 0.76 | 0.37 | 2.08 |
|  | Pre-Pandemic-#Wholesale and Retail Trade Motor Repair | 1.12 | 0.43 | 0.29 | 0.78 | 0.52 | 2.38 |
|  | Post-Pandemic-#Wholesale and Retail Trade Motor Repair | 1.23 | 0.38 | 0.67 | 0.51 | 0.67 | 2.25 |
|  | Pre-Pandemic-#Transportation and Storage | 0.39 | 0.21 | -1.75 | 0.08 | 0.13 | 1.12 |
|  | Post-Pandemic-#Transportation and Storage | 0.92 | 0.40 | -0.20 | 0.84 | 0.39 | 2.15 |
|  | Pre-Pandemic-#Accommodation and Food Services | 0.71 | 0.44 | -0.56 | 0.58 | 0.21 | 2.41 |
|  | Post-Pandemic-#Accommodation and Food Services | 1.38 | 0.51 | 0.87 | 0.38 | 0.67 | 2.87 |
|  | Pre-Pandemic-#Information and Communication | 1.30 | 0.77 | 0.44 | 0.66 | 0.40 | 4.16 |
|  | Post-Pandemic-#Information and Communication | 1.57 | 0.61 | 1.17 | 0.24 | 0.74 | 3.35 |
|  | Pre-Pandemic-#Financial and Insurance Activities | 0.38 | 0.16 | -2.28 | 0.02 | 0.16 | 0.87 |
|  | Post-Pandemic-#Financial and Insurance Activities | 0.75 | 0.25 | -0.84 | 0.40 | 0.39 | 1.46 |
|  | Pre-Pandemic-#Real Estate Activities | 2.50 | 1.29 | 1.78 | 0.08 | 0.91 | 6.89 |
|  | Post-Pandemic-#Real Estate Activities | 0.96 | 0.58 | -0.07 | 0.94 | 0.29 | 3.16 |
|  | Pre-Pandemic-#Professional Scientific and Technical | 0.76 | 0.35 | -0.58 | 0.56 | 0.31 | 1.90 |
|  | Post-Pandemic-#Professional Scientific and Technical | 1.53 | 0.44 | 1.48 | 0.14 | 0.87 | 2.67 |
|  | Pre-Pandemic-#Administrative and Support Services | 0.24 | 0.17 | -2.03 | 0.04 | 0.06 | 0.95 |
|  | Post-Pandemic-#Administrative and Support Services | 1.03 | 0.31 | 0.09 | 0.93 | 0.57 | 1.85 |
|  | Pre-Pandemic-#Public Administration and Defence | 0.78 | 0.22 | -0.89 | 0.37 | 0.45 | 1.35 |
|  | Post-Pandemic-#Public Administration and Defence | 0.95 | 0.24 | -0.19 | 0.85 | 0.58 | 1.57 |
|  | Pre-Pandemic-#Education | 0.85 | 0.22 | -0.63 | 0.53 | 0.51 | 1.41 |
|  | Post-Pandemic-#Education | 1.15 | 0.27 | 0.57 | 0.57 | 0.72 | 1.83 |
|  | Post-Pandemic-#Human Health and Social Work Activities | 1.25 | 0.28 | 0.96 | 0.34 | 0.80 | 1.95 |
|  | Pre-Pandemic-#Other Services | 0.26 | 0.12 | -3.01 | 0.00 | 0.11 | 0.62 |
|  | Post-Pandemic-#Other Services | 0.74 | 0.23 | -0.95 | 0.34 | 0.41 | 1.37 |
| Union membership: yes | | 0.89 | 0.20 | -0.55 | 0.58 | 0.57 | 1.37 |
| Pre-Post Pandemic # Industry # Union membership | |  |  |  |  |  |  |
|  | Pre-Pandemic-#Mining, Energy and Water Supply#Yes | 1.75 | 1.72 | 0.57 | 0.57 | 0.26 | 12.00 |
|  | Post-Pandemic-#Mining, Energy and Water Supply#Yes | 1.04 | 0.74 | 0.05 | 0.96 | 0.26 | 4.17 |
|  | Pre-Pandemic-#Manufacturing#Yes | 3.19 | 2.18 | 1.69 | 0.09 | 0.83 | 12.19 |
|  | Post-Pandemic-#Manufacturing#Yes | 1.36 | 0.70 | 0.60 | 0.55 | 0.50 | 3.74 |
|  | Pre-Pandemic-#Construction#Yes | 0.71 | 0.65 | -0.38 | 0.71 | 0.12 | 4.29 |
|  | Post-Pandemic-#Construction#Yes | 1.41 | 0.85 | 0.57 | 0.57 | 0.43 | 4.60 |
|  | Pre-Pandemic-#Wholesale and Retail Trade Motor Repair#Yes | 0.92 | 0.47 | -0.17 | 0.87 | 0.33 | 2.52 |
|  | Post-Pandemic-#Wholesale and Retail Trade Motor Repair#Yes | 0.90 | 0.36 | -0.27 | 0.79 | 0.41 | 1.98 |
|  | Pre-Pandemic-#Transportation and Storage#Yes | 2.60 | 1.65 | 1.50 | 0.13 | 0.75 | 9.01 |
|  | Post-Pandemic-#Transportation and Storage#Yes | 1.65 | 0.84 | 0.99 | 0.32 | 0.61 | 4.45 |
|  | Pre-Pandemic-#Accommodation and Food Services#Yes | 0.07 | 0.09 | -2.13 | 0.03 | 0.01 | 0.80 |
|  | Post-Pandemic-#Accommodation and Food Services#Yes | 0.92 | 0.61 | -0.12 | 0.91 | 0.25 | 3.39 |
|  | Pre-Pandemic-#Information and Communication#Yes | 2.06 | 1.79 | 0.83 | 0.41 | 0.37 | 11.30 |
|  | Post-Pandemic-#Information and Communication#Yes | 0.50 | 0.26 | -1.34 | 0.18 | 0.18 | 1.38 |
|  | Pre-Pandemic-#Financial and Insurance Activities#Yes | 3.78 | 2.56 | 1.97 | 0.05 | 1.00 | 14.22 |
|  | Post-Pandemic-#Financial and Insurance Activities#Yes | 0.72 | 0.39 | -0.61 | 0.54 | 0.25 | 2.07 |
|  | Pre-Pandemic-#Real Estate Activities#Yes | 1.80 | 1.48 | 0.72 | 0.47 | 0.36 | 8.99 |
|  | Post-Pandemic-#Real Estate Activities#Yes | 0.51 | 0.38 | -0.91 | 0.36 | 0.12 | 2.17 |
|  | Pre-Pandemic-#Professional Scientific and Technical#Yes | 1.13 | 0.72 | 0.19 | 0.85 | 0.32 | 3.97 |
|  | Post-Pandemic-#Professional Scientific and Technical#Yes | 0.84 | 0.30 | -0.50 | 0.62 | 0.42 | 1.68 |
|  | Pre-Pandemic-#Administrative and Support Services#Yes | 8.34 | 8.07 | 2.19 | 0.03 | 1.25 | 55.62 |
|  | Post-Pandemic-#Administrative and Support Services#Yes | 0.74 | 0.31 | -0.72 | 0.47 | 0.33 | 1.67 |
|  | Pre-Pandemic-#Public Administration and Defence#Yes | 1.34 | 0.47 | 0.85 | 0.40 | 0.68 | 2.66 |
|  | Post-Pandemic-#Public Administration and Defence#Yes | 1.49 | 0.45 | 1.32 | 0.19 | 0.83 | 2.67 |
|  | Pre-Pandemic-#Education#Yes | 1.13 | 0.36 | 0.37 | 0.71 | 0.60 | 2.12 |
|  | Post-Pandemic-#Education#Yes | 1.17 | 0.33 | 0.57 | 0.57 | 0.68 | 2.02 |
|  | Post-Pandemic-#Human Health and Social Work Activities#Yes | 1.07 | 0.29 | 0.25 | 0.80 | 0.63 | 1.80 |
|  | Pre-Pandemic-#Other Services #Yes | 8.86 | 5.47 | 3.53 | 0.00 | 2.64 | 29.74 |
|  | Post-Pandemic-#Other Services #Yes | 1.54 | 0.57 | 1.16 | 0.25 | 0.74 | 3.17 |
| Country: | |  |  |  |  |  |  |
|  | Wales | 1.04 | 0.17 | 0.26 | 0.79 | 0.76 | 1.44 |
|  | Scotland | 0.95 | 0.13 | -0.36 | 0.72 | 0.74 | 1.23 |
|  | Northern Ireland | 0.90 | 0.16 | -0.60 | 0.55 | 0.63 | 1.28 |
| Highest level of education: | |  |  |  |  |  |  |
|  | A-Level | 0.75 | 0.08 | -2.68 | 0.01 | 0.61 | 0.92 |
|  | GCSE | 0.64 | 0.08 | -3.64 | 0.00 | 0.50 | 0.81 |
|  | Other | 0.45 | 0.11 | -3.34 | 0.00 | 0.28 | 0.72 |
|  | None | 0.42 | 0.19 | -1.96 | 0.05 | 0.17 | 1.00 |
| Self-reported financial conditions: bad | | 2.75 | 0.23 | 12.24 | 0.00 | 2.34 | 3.23 |
| Health condition: yes | | 1.92 | 0.17 | 7.49 | 0.00 | 1.62 | 2.27 |
| Company size: | |  |  |  |  |  |  |
|  | 25-199 | 0.94 | 0.11 | -0.53 | 0.60 | 0.75 | 1.18 |
|  | More than 200 | 1.05 | 0.12 | 0.43 | 0.67 | 0.84 | 1.31 |
| Wave Number | |  |  |  |  |  |  |
|  | Wave 10 | 0.88 | 0.09 | -1.18 | 0.24 | 0.72 | 1.09 |
|  | Wave 11 | 1.35 | 0.16 | 2.55 | 0.01 | 1.07 | 1.70 |
|  | **COVID-19 sweeps** |  |  |  |  |  |  |
|  | Apr-20 | 2.59 | 0.28 | 8.70 | 0.00 | 2.09 | 3.21 |
|  | May-20 | 1.79 | 0.20 | 5.06 | 0.00 | 1.43 | 2.24 |
|  | Jun-20 | 1.55 | 0.19 | 3.52 | 0.00 | 1.21 | 1.97 |
|  | Jul-20 | 0.87 | 0.10 | -1.20 | 0.23 | 0.69 | 1.09 |
|  | Sep-20 | 1.12 | 0.13 | 0.98 | 0.33 | 0.89 | 1.42 |
|  | Nov-20 | 1.95 | 0.24 | 5.31 | 0.00 | 1.52 | 2.49 |
|  | Jan-21 | 2.23 | 0.25 | 7.01 | 0.00 | 1.78 | 2.79 |
|  | Mar-21 | 1.27 | 0.16 | 1.90 | 0.06 | 0.99 | 1.62 |
|  | Sep-21 | 1.00 | (omitted) |  |  |  |  |
| Constant | | 0.03 | 0.02 | -5.33 | 0.00 | 0.01 | 0.10 |

Note: Observations=27,971; Groups=3,341 (Minimum=1; Average= 8.4; Maximum=12).

### Supplementary file S3. Sample characteristics for trade union presence and membership

#### Table S3.1. Sample characteristics for union presence in the pre and pandemic USoc Waves*

|  |  | **Trade Union presence**** | | | **Trade Union membership**** | | |
| --- | --- | --- | --- | --- | --- | --- | --- |
|  |  | **Pre-Pandemic** | **Pandemic** | **Total** | **Pre-Pandemic** | **Pandemic** | **Total** |
| **Outcome** | *GHQ case* |  |  |  |  |  |  |
|  | No | 11,636 | 27,254 | 38,890 | 6,078 | 15,510 | 21,588 |
|  | % | 82.3 (81.6) | 76.2 (74.6) | 77.9 (76.5) | 81.4 (80.6) | 75.6 (74.7) | 77.2 (76.2) |
|  | Yes | 2,506 | 8,519 | 11,025 | 1,389 | 4,994 | 6,383 |
|  | % | 17.7 (18.4) | 23.8 (25.4) | 22.1 (23.5) | 18.6 (19.4) | 24.4 (25.3) | 22.8 (23.8) |
|  | **Total** | **14,142** | **35,773** | **49,915** | **7,467** | **20,504** | **27,971** |
|  | **%** | **100.0** | **100.0** | **100.0** | **100.0** | **100.0** | **100.0** |
| **Confounders** | *Sex* |  |  |  |  |  |  |
|  | Male | 5,815 | 14,225 | 20,040 | 2,672 | 7,081 | 9,753 |
|  | % | 41.1 (44.1) | 39.8 (46.3) | 40.1 (45.7) | 35.8 (38.9) | 34.5 (41.5) | 34.9 (40.8) |
|  | Female | 8,327 | 21,548 | 29,875 | 4,795 | 13,423 | 18,218 |
|  | % | 58.9 (55.9) | 60.2 (53.7) | 59.9 (54.3) | 64.2 (61.1) | 65.5 (58.5) | 65.1 (59.2) |
|  | *Race* |  |  |  |  |  |  |
|  | White | 13,040 | 33,063 | 46,103 | 6,865 | 18,924 | 25,789 |
|  | % | 92.2 (94.6) | 92.4 (94.4) | 92.4 (94.5) | 91.9 (94.8) | 92.3 (94.7) | 92.2 (94.7) |
|  | Non-white | 1,102 | 2,710 | 3,812 | 602 | 1,580 | 2,182 |
|  | % | 7.8 (5.4) | 7.6 (5.6) | 7.6 (5.5) | 8.1 (5.2) | 7.7 (5.3) | 7.8 (5.3) |
|  | *UK country* |  |  |  |  |  |  |
|  | England | 11,238 | 28,932 | 40,170 | 5,715 | 16,143 | 21,858 |
|  | % | 79.5 (85.7) | 80.9 (85.2) | 80.5 (85.3) | 76.5 (83.4) | 78.7 (83.7) | 78.1 (83.6) |
|  | Wales | 926 | 2,165 | 3,091 | 571 | 1,408 | 1,979 |
|  | % | 6.5 (4.5) | 6.1 (4.7) | 6.2 (4.7) | 7.6 (5.3) | 6.9 (5.1) | 7.1 (5.2) |
|  | Scotland | 1,278 | 3,204 | 4,482 | 763 | 2,007 | 2,770 |
|  | % | 9.0 (7.8) | 9.0 (7.7) | 9.0 (7.7) | 10.2 (9.1) | 9.8 (8.9) | 9.9 (8.9) |
|  | Northern Ireland | 700 | 1,472 | 2,172 | 418 | 946 | 1,364 |
|  | % | 4.9 (2.0) | 4.1 (2.4) | 4.4 (2.3) | 5.6 (2.2) | 4.6 (2.3) | 4.9 (2.3) |
|  | *Qualifications* |  |  |  |  |  |  |
|  | Degree | 8,110 | 20,756 | 28,866 | 4,562 | 12,540 | 17,102 |
|  | % | 57.3 (56.4) | 58.0 (51.6) | 57.8 (52.9) | 61.1 (59.7) | 61.2 (55.1) | 61.1 (56.3) |
|  | A-Level | 2,904 | 7,376 | 10,280 | 1,470 | 4,043 | 5,513 |
|  | % | 20.5 (20.8) | 20.6 (23.0) | 20.6 (22.5) | 19.7 (19.8) | 19.7 (21.1) | 19.7 (20.8) |
|  | GCSE | 2,347 | 5,848 | 8,195 | 1,095 | 3,046 | 4,141 |
|  | % | 16.6 (17.0) | 16.3 (19.5) | 16.4 (18.8) | 14.7 (15.6) | 14.9 (18.8) | 14.8 (18.0) |
|  | Other | 596 | 1,403 | 1,999 | 259 | 668 | 927 |
|  | % | 4.2 (4.5) | 3.9 (4.6) | 4.0 (4.6) | 3.5 (3.8) | 3.3 (3.8) | 3.3 (3.8) |
|  | None | 185 | 390 | 575 | 81 | 207 | 288 |
|  | % | 1.3 (1.3) | 1.1 (1.2) | 1.2 (1.2) | 1.1 (1.2) | 1.0 (1.2) | 1.0 (1.2) |
|  | *Financial situation* |  |  |  |  |  |  |
|  | Good | 10,652 | 28,892 | 39,544 | 5,580 | 16,788 | 22,368 |
|  | % | 75.3 (74.4) | 80.8 (77.3) | 79.2 (76.5) | 74.7 (73.5) | 81.9 (78.6) | 80.0 (77.3) |
|  | Bad | 3,490 | 6,881 | 10,371 | 1,887 | 3,716 | 5,603 |
|  | % | 24.7 (25.6) | 19.2 (22.7) | 20.8 (23.5) | 25.3 (26.5) | 18.1 (21.4) | 20.0 (22.7) |
|  | *Past health condition* |  |  |  |  |  |  |
|  | No | 10,325 | 25,776 | 36,101 | 5,377 | 14,537 | 19,914 |
|  | % | 73.0 (72.6) | 72.1 (71.6) | 72.3 (71.9) | 72.0 (71.4) | 70.9 (70.9) | 71.2 (71.0) |
|  | Yes | 3,817 | 9,997 | 13,814 | 2,090 | 5,967 | 8,057 |
|  | % | 27.0 (27.4) | 27.9 (28.4) | 27.7 (28.2) | 28.0 (28.6) | 29.1 (29.1) | 28.8 (29.0) |
|  | *Job size* |  |  |  |  |  |  |
|  | 1 to 24 employees | 3,701 | 9,197 | 12,898 | 1,059 | 3,248 | 4,307 |
|  | % | 26.2 (26.6) | 25.7 (27.4) | 25.8 (27.2) | 26.2 (14.5) | 25.7 (15.9) | 25.8 (15.6) |
|  | 25 to 199 employees | 5,301 | 12,724 | 18,025 | 2,855 | 7,358 | 10,213 |
|  | % | 37.5 (37.8) | 35.6 (36.3) | 36.1 (36.7) | 37.5 (38.0) | 35.6 (36.9) | 36.1 (37.2) |
|  | More than 200 employees | 5,140 | 13,852 | 18,992 | 3,553 | 9,898 | 13,451 |
|  | % | 36.4 (35.6) | 38.7 (36.4) | 38.1 (36.2) | 36.4 (47.5) | 38.7 (47.2) | 38.1 (47.3) |
|  | **Total** | **14,142** | **35,773** | **49,915** | **7,467** | **20,504** | **27,971** |
|  | **%** | **100.0** | **100.0** | **100.0** | **100.0** | **100.0** | **100.0** |
|  | *Age (continuous)* | **Mean**  **sd** | **Mean**  **sd** | **Mean**  **sd** | **Mean**  **sd** | **Mean**  **sd** | **Mean**  **sd** |
|  |  | 45.7 (44.59)  11.3 (11.8) | 47.7 (44.9)  11.3 (12.3) | 47.2 (44.8)  11.4 (12.2) | 46.1 (45.2)  10.6 (10.8) | 48.2 (45.9)  10.8 (11.7) | 47.7 (45.8)  10.8 (11.6) |

*Calculation using the weighted Sample (in brackets))

#### Table S3.2. Observations for Industry (SIC-2007) for union presence in pre-pandemic and pandemic Waves (Unweighted and Weighted Sample (in brackets))

| **Industry SIC-2007** | **Trade Union presence** | | | **Trade Union membership** | | |
| --- | --- | --- | --- | --- | --- | --- |
|  | Pre-pandemic | Pandemic | Total | Pre-pandemic | Pandemic | Total |
| Mining, Energy and Water Supply | 210 | 727 | 937 | 118 | 348 | 466 |
| % Unweighted (Weighted) | 1.5(1.4) | 2.0(2.2) | 1.9(1.9) | 1.58(1.4) | 1.73(1.9) | 1.68(1.8) |
| Manufacturing | 1,319 | 2,722 | 4,041 | 381 | 866 | 1,247 |
| % Unweighted (Weighted) | 9.3(9.3) | 7.6(8.0) | 8.1(8.4) | 5.16(5.0) | 4.17(4.8) | 4.43(4.9) |
| Construction | 456 | 1,252 | 1,734 | 99 | 304 | 403 |
| % Unweighted (Weighted) | 3.2(3.4) | 3.5(4.6) | 3.4(4.3) | 1.3(1.4) | 1.5(2.0) | 1.5(1.8) |
| Wholesale and Retail Trade Motor Repair | 1,599 | 3,680 | 5,279 | 532 | 1,397 | 1,929 |
| % Unweighted (Weighted) | 11.3(12.4) | 10.3(12.6) | 10.6(12.6) | 7.2(8.7) | 6.9(9.2) | 7.0(9.0) |
| Transportation and Storage | 574 | 1,224 | 1,798 | 340 | 775 | 1,115 |
| % Unweighted (Weighted) | 4.1(4.1) | 3.4(4.0) | 3.6(4.0) | 4.6(4.6) | 3.8(4.7) | 4.0(4.7) |
| Accommodation and Food Services | 393 | 800 | 1,193 | 78 | 198 | 276 |
| % Unweighted (Weighted) | 2.8(3.0) | 2.2(2.7) | 2.4(2.8) | 1.1(1.1) | 1.0(1.1) | 1.0(1.1) |
| Information and Communication | 574 | 1,616 | 2,190 | 154 | 575 | 729 |
| % Unweighted (Weighted) | 4.1(4.1) | 4.5(4.5) | 4.4(4.4) | 2.1(2.4) | 2.8(3.1) | 2.6(2.9) |
| Financial and Insurance Activities | 537 | 1,749 | 2,286 | 224 | 774 | 998 |
| % Unweighted (Weighted) | 3.8(4.1) | 4.9(4.5) | 4.6(4.4) | 3.0(3.1) | 3.8(3.5) | 3.6(3.4) |
| Real Estate Activities | 164 | 329 | 493 | 66 | 102 | 168 |
| % Unweighted (Weighted) | 1.2(1.2) | 0.9(0.8) | 1.0(0.9) | 0.9(1.0) | 0.5(0.5) | 0.6(0.6) |
| Professional Scientific and Technical | 970 | 2,260 | 3,230 | 197 | 830 | 1,027 |
| % Unweighted (Weighted) | 6.9(6.9) | 6.3(5.4) | 6.5(5.8) | 2.6(2.5) | 4.1(3.7) | 3.7(3.4) |
| Administrative and Support Services | 445 | 1,500 | 1,945 | 126 | 887 | 1,013 |
| % Unweighted (Weighted) | 3.2(3.5) | 4.2(4.0) | 3.9(3.9) | 1.7(1.9) | 4.3(4.5) | 3.6(3.8) |
| Public Administration and Defence | 1,426 | 2,575 | 4,001 | 1,282 | 2,326 | 3,608 |
| % Unweighted (Weighted) | 10.1(9.2) | 7.2(6.5) | 8.0(7.2) | 17.0(16.1) | 11.2(10.3) | 12.8(11.8) |
| Education | 2,250 | 6,024 | 8,274 | 1,877 | 5,136 | 7,013 |
| % Unweighted (Weighted) | 15.9(15.5) | 16.8(15.1) | 16.6(15.2) | 25.2(25.1) | 25.1(23.4) | 25.2(23.8) |
| Human Health and Social Work Activities | 2,717 | 6,253 | 8,970 | 1,803 | 4,567 | 6,370 |
| % Unweighted (Weighted) | 19.2(18.1) | 17.5(15.9) | 18.0(16.5) | 24.2(23.0) | 22.1(19.5) | 22.7(20.4) |
| Other Services | 508 | 3,062 | 3,570 | 190 | 1,419 | 1,609 |
| % Unweighted (Weighted) | 3.6(3.8) | 8.6(9.5) | 7.2(7.9) | 2.5(2.8) | 7.1(7.9) | 5.8(6.6) |
| **Total** | **14,142** | **35,773** | **49,915** | **7,467** | **20,504** | **27,971** |
| **%** | **100.00** | **100.00** | **100.00** | **100.00** | **100.00** | **100.00** |

### Supplementary file S4. Sensitivity analyses

#### Union presence – adjusted model (2-way interaction), excluding union members

|  |  | **Adjusted model (weighted)** | | | | | |
| --- | --- | --- | --- | --- | --- | --- | --- |
|  |  | **OR** | **std. err.** | **z** | **P>z** | **[95% conf.** | **interval]** |
| sex |  | 2.49 | 0.20 | 11.23 | 0.00 | 2.12 | 2.92 |
| age |  | 1.02 | 0.02 | 0.84 | 0.40 | 0.98 | 1.06 |
| age_square | | 1.00 | 0.00 | -2.22 | 0.03 | 1.00 | 1.00 |
| Ethnicity | | 0.88 | 0.13 | -0.87 | 0.38 | 0.66 | 1.17 |
| Pre-Post Pandemic | | 1.53 | 0.20 | 3.30 | 0.00 | 1.19 | 1.98 |
| Union presence: yes | | 0.97 | 0.11 | -0.30 | 0.76 | 0.77 | 1.21 |
| Pre-Post Pandemic # Union presence: yes | | 0.94 | 0.12 | -0.47 | 0.64 | 0.74 | 1.20 |
| Industry: | |  |  |  |  |  |  |
|  | Mining, Energy and Water Supply | 1.18 | 0.35 | 0.55 | 0.58 | 0.66 | 2.10 |
|  | Manufacturing | 0.65 | 0.14 | -2.01 | 0.04 | 0.43 | 0.99 |
|  | Construction | 0.63 | 0.15 | -1.91 | 0.06 | 0.39 | 1.01 |
|  | Wholesale and Retail Trade Motor Repair | 0.84 | 0.17 | -0.83 | 0.40 | 0.57 | 1.26 |
|  | Transportation and Storage | 0.53 | 0.14 | -2.37 | 0.02 | 0.31 | 0.89 |
|  | Information and Communication | 0.85 | 0.19 | -0.72 | 0.47 | 0.55 | 1.32 |
|  | Financial and Insurance Activities | 0.64 | 0.16 | -1.82 | 0.07 | 0.40 | 1.03 |
|  | Real Estate Activities | 0.98 | 0.30 | -0.05 | 0.96 | 0.54 | 1.79 |
|  | Professional Scientific and Technical | 0.76 | 0.16 | -1.34 | 0.18 | 0.51 | 1.13 |
|  | Administrative and Support Services | 0.59 | 0.14 | -2.30 | 0.02 | 0.38 | 0.93 |
|  | Public Administration and Defence | 0.65 | 0.15 | -1.92 | 0.06 | 0.42 | 1.01 |
|  | Education | 0.74 | 0.15 | -1.50 | 0.13 | 0.49 | 1.10 |
|  | Human Health and Social Work Activities | 0.75 | 0.15 | -1.43 | 0.15 | 0.51 | 1.11 |
|  | Other Services | 0.62 | 0.13 | -2.32 | 0.02 | 0.41 | 0.93 |
| Country: | |  |  |  |  |  |  |
|  | Wales | 1.43 | 0.23 | 2.21 | 0.03 | 1.04 | 1.97 |
|  | Scotland | 1.04 | 0.15 | 0.28 | 0.78 | 0.78 | 1.39 |
|  | Northern Ireland | 0.91 | 0.18 | -0.50 | 0.61 | 0.62 | 1.33 |
| Highest level of education: | |  |  |  |  |  |  |
|  | A-Level | 0.70 | 0.07 | -3.82 | 0.00 | 0.58 | 0.84 |
|  | GCSE | 0.82 | 0.09 | -1.85 | 0.06 | 0.67 | 1.01 |
|  | Other | 0.60 | 0.12 | -2.45 | 0.01 | 0.40 | 0.90 |
|  | None | 0.35 | 0.12 | -2.98 | 0.00 | 0.17 | 0.70 |
| Self-reported financial conditions: bad | | 2.27 | 0.19 | 9.69 | 0.00 | 1.92 | 2.68 |
| Health condition: yes | | 2.16 | 0.18 | 9.44 | 0.00 | 1.84 | 2.53 |
| Company size: | |  |  |  |  |  |  |
|  | 25-199 | 0.87 | 0.08 | -1.46 | 0.14 | 0.72 | 1.05 |
|  | More than 200 | 0.96 | 0.09 | -0.47 | 0.64 | 0.79 | 1.15 |
| Wave Number | |  |  |  |  |  |  |
|  | Wave 10 | 0.92 | 0.09 | -0.80 | 0.42 | 0.76 | 1.12 |
|  | Wave 11 | 1.31 | 0.14 | 2.53 | 0.01 | 1.06 | 1.63 |
|  | COVID-19 Sweeps |  |  |  |  |  |  |
|  | Apr-20 | 2.23 | 0.25 | 7.14 | 0.00 | 1.79 | 2.78 |
|  | May-20 | 1.83 | 0.21 | 5.22 | 0.00 | 1.46 | 2.29 |
|  | Jun-20 | 1.69 | 0.20 | 4.31 | 0.00 | 1.33 | 2.14 |
|  | Jul-20 | 1.01 | 0.12 | 0.10 | 0.92 | 0.81 | 1.26 |
|  | Sep-20 | 0.96 | 0.12 | -0.36 | 0.72 | 0.75 | 1.22 |
|  | Nov-20 | 1.89 | 0.24 | 5.00 | 0.00 | 1.47 | 2.43 |
|  | Jan-21 | 1.63 | 0.20 | 4.07 | 0.00 | 1.29 | 2.07 |
|  | Mar-21 | 1.27 | 0.15 | 1.98 | 0.05 | 1.00 | 1.61 |
|  | Sep-21 | 1.00 | (omitted) |  |  |  |  |
| Constant |  | 0.03 | 0.02 | -6.77 | 0.00 | 0.01 | 0.08 |

Note: Observations=39,190; Groups=4,405 (Minimum=1; Average= 7.8; Maximum=12).

#### Union presence – adjusted model (3-way interaction), excluding union members

|  |  | **Adjusted model (weighted) -- three-ways interaction** | | | | | |
| --- | --- | --- | --- | --- | --- | --- | --- |
|  |  | **OR** | **std. err.** | **z** | **P>z** | **[95% conf.** | **interval]** |
| Sex |  | 2.48 | 0.20 | 11.13 | 0.00 | 2.11 | 2.91 |
| Age | | 1.02 | 0.02 | 0.94 | 0.35 | 0.98 | 1.06 |
| Age square | | 1.00 | 0.00 | -2.33 | 0.02 | 1.00 | 1.00 |
| Ethniciy | | 0.87 | 0.13 | -0.98 | 0.33 | 0.65 | 1.15 |
| Pre-Post Pandemic # Industry: |  |  |  |  |  |  |  |
|  | Pre-Pandemic-#Mining, Energy and Water Supply | 3.04 | 1.73 | 1.95 | 0.05 | 0.99 | 9.29 |
|  | Post-Pandemic-#Mining, Energy and Water Supply | 2.31 | 0.74 | 2.62 | 0.01 | 1.23 | 4.34 |
|  | Pre-Pandemic-#Manufacturing | 0.85 | 0.21 | -0.67 | 0.51 | 0.52 | 1.38 |
|  | Post-Pandemic-#Manufacturing | 1.46 | 0.37 | 1.51 | 0.13 | 0.89 | 2.40 |
|  | Pre-Pandemic-#Construction | 0.72 | 0.23 | -1.03 | 0.31 | 0.38 | 1.35 |
|  | Post-Pandemic-#Construction | 1.25 | 0.34 | 0.82 | 0.41 | 0.73 | 2.15 |
|  | Pre-Pandemic-#Wholesale and Retail Trade Motor Repair | 1.11 | 0.28 | 0.40 | 0.69 | 0.67 | 1.83 |
|  | Post-Pandemic-#Wholesale and Retail Trade Motor Repair | 1.61 | 0.42 | 1.81 | 0.07 | 0.96 | 2.68 |
|  | Pre-Pandemic-#Transportation and Storage | 1.00 | 0.38 | 0.01 | 0.99 | 0.48 | 2.10 |
|  | Post-Pandemic-#Transportation and Storage | 0.91 | 0.32 | -0.27 | 0.79 | 0.46 | 1.81 |
|  | Pre-Pandemic-#Accommodation and Food Services | 1.61 | 0.50 | 1.51 | 0.13 | 0.87 | 2.97 |
|  | Post-Pandemic-#Accommodation and Food Services | 1.81 | 0.56 | 1.94 | 0.05 | 0.99 | 3.31 |
|  | Pre-Pandemic-#Information and Communication | 0.75 | 0.27 | -0.79 | 0.43 | 0.37 | 1.52 |
|  | Post-Pandemic-#Information and Communication | 1.74 | 0.42 | 2.27 | 0.02 | 1.08 | 2.80 |
|  | Pre-Pandemic-#Financial and Insurance Activities | 1.49 | 0.46 | 1.31 | 0.19 | 0.82 | 2.73 |
|  | Post-Pandemic-#Financial and Insurance Activities | 1.11 | 0.30 | 0.37 | 0.71 | 0.65 | 1.88 |
|  | Pre-Pandemic-#Real Estate Activities | 1.07 | 0.47 | 0.16 | 0.87 | 0.45 | 2.55 |
|  | Post-Pandemic-#Real Estate Activities | 1.78 | 0.72 | 1.41 | 0.16 | 0.80 | 3.95 |
|  | Pre-Pandemic-#Professional Scientific and Technical | 0.87 | 0.20 | -0.62 | 0.54 | 0.55 | 1.37 |
|  | Post-Pandemic-#Professional Scientific and Technical | 1.47 | 0.34 | 1.66 | 0.10 | 0.93 | 2.32 |
|  | Pre-Pandemic-#Administrative and Support Services | 0.59 | 0.21 | -1.47 | 0.14 | 0.30 | 1.19 |
|  | Post-Pandemic-#Administrative and Support Services | 1.14 | 0.31 | 0.47 | 0.64 | 0.66 | 1.96 |
|  | Pre-Pandemic-#Public Administration and Defence | 0.44 | 0.27 | -1.35 | 0.18 | 0.13 | 1.46 |
|  | Post-Pandemic-#Public Administration and Defence | 1.22 | 0.39 | 0.61 | 0.54 | 0.65 | 2.29 |
|  | Pre-Pandemic-#Education | 0.46 | 0.16 | -2.29 | 0.02 | 0.24 | 0.90 |
|  | Post-Pandemic-#Education | 1.67 | 0.44 | 1.95 | 0.05 | 1.00 | 2.79 |
|  | Post-Pandemic-#Human Health and Social Work Activities | 1.33 | 0.29 | 1.31 | 0.19 | 0.87 | 2.04 |
|  | Pre-Pandemic-#Other Services | 1.17 | 0.38 | 0.49 | 0.63 | 0.62 | 2.20 |
|  | Post-Pandemic-#Other Services | 1.30 | 0.29 | 1.17 | 0.24 | 0.84 | 2.02 |
| Union presence: yes | | 1.13 | 0.27 | 0.50 | 0.62 | 0.70 | 1.81 |
| Pre-Post Pandemic # Industry # Union presence | |  |  |  |  |  |  |
|  | Pre-Pandemic-#Mining, Energy and Water Supply#Yes | 0.35 | 0.35 | -1.06 | 0.29 | 0.05 | 2.43 |
|  | Post-Pandemic-#Mining, Energy and Water Supply#Yes | 0.53 | 0.40 | -0.84 | 0.40 | 0.12 | 2.34 |
|  | Pre-Pandemic-#Manufacturing#Yes | 0.31 | 0.19 | -1.90 | 0.06 | 0.09 | 1.04 |
|  | Post-Pandemic-#Manufacturing#Yes | 0.34 | 0.18 | -2.08 | 0.04 | 0.13 | 0.94 |
|  | Pre-Pandemic-#Construction#Yes | 1.88 | 1.34 | 0.89 | 0.37 | 0.47 | 7.57 |
|  | Post-Pandemic-#Construction#Yes | 0.50 | 0.27 | -1.28 | 0.20 | 0.18 | 1.44 |
|  | Pre-Pandemic-#Wholesale and Retail Trade Motor Repair#Yes | 1.00 | 0.44 | -0.01 | 0.99 | 0.42 | 2.39 |
|  | Post-Pandemic-#Wholesale and Retail Trade Motor Repair#Yes | 0.82 | 0.32 | -0.50 | 0.62 | 0.38 | 1.78 |
|  | Pre-Pandemic-#Transportation and Storage#Yes | 0.36 | 0.22 | -1.70 | 0.09 | 0.11 | 1.17 |
|  | Post-Pandemic-#Transportation and Storage#Yes | 0.95 | 0.55 | -0.09 | 0.93 | 0.31 | 2.93 |
|  | Pre-Pandemic-#Accommodation and Food Services#Yes | 0.45 | 0.31 | -1.16 | 0.25 | 0.12 | 1.72 |
|  | Post-Pandemic-#Accommodation and Food Services#Yes | 0.80 | 0.40 | -0.45 | 0.66 | 0.30 | 2.13 |
|  | Pre-Pandemic-#Information and Communication#Yes | 1.84 | 1.24 | 0.90 | 0.37 | 0.49 | 6.92 |
|  | Post-Pandemic-#Information and Communication#Yes | 0.94 | 0.43 | -0.14 | 0.89 | 0.38 | 2.30 |
|  | Pre-Pandemic-#Financial and Insurance Activities#Yes | 0.33 | 0.16 | -2.22 | 0.03 | 0.13 | 0.88 |
|  | Post-Pandemic-#Financial and Insurance Activities#Yes | 0.89 | 0.37 | -0.29 | 0.77 | 0.39 | 2.00 |
|  | Pre-Pandemic-#Real Estate Activities#Yes | 2.75 | 2.02 | 1.38 | 0.17 | 0.65 | 11.61 |
|  | Post-Pandemic-#Real Estate Activities#Yes | 0.54 | 0.47 | -0.71 | 0.48 | 0.10 | 2.94 |
|  | Pre-Pandemic-#Professional Scientific and Technical#Yes | 0.83 | 0.43 | -0.35 | 0.72 | 0.31 | 2.27 |
|  | Post-Pandemic-#Professional Scientific and Technical#Yes | 1.03 | 0.36 | 0.08 | 0.94 | 0.52 | 2.05 |
|  | Pre-Pandemic-#Administrative and Support Services#Yes | 0.40 | 0.32 | -1.16 | 0.25 | 0.08 | 1.88 |
|  | Post-Pandemic-#Administrative and Support Services#Yes | 1.04 | 0.40 | 0.09 | 0.93 | 0.48 | 2.22 |
|  | Pre-Pandemic-#Public Administration and Defence#Yes | 1.83 | 1.24 | 0.89 | 0.37 | 0.48 | 6.88 |
|  | Post-Pandemic-#Public Administration and Defence#Yes | 0.88 | 0.34 | -0.31 | 0.75 | 0.41 | 1.90 |
|  | Pre-Pandemic-#Education#Yes | 1.85 | 0.77 | 1.48 | 0.14 | 0.82 | 4.17 |
|  | Post-Pandemic-#Education#Yes | 0.73 | 0.24 | -0.94 | 0.35 | 0.39 | 1.40 |
|  | Post-Pandemic-#Human Health and Social Work Activities#Yes | 0.98 | 0.28 | -0.07 | 0.94 | 0.56 | 1.73 |
|  | Pre-Pandemic-#Other Services #Yes | 0.20 | 0.11 | -2.87 | 0.00 | 0.07 | 0.60 |
|  | Post-Pandemic-#Other Services #Yes | 0.58 | 0.22 | -1.44 | 0.15 | 0.28 | 1.21 |
| Country: | |  |  |  |  |  |  |
|  | Wales | 1.44 | 0.24 | 2.21 | 0.03 | 1.04 | 1.98 |
|  | Scotland | 1.03 | 0.15 | 0.17 | 0.87 | 0.76 | 1.38 |
|  | Northern Ireland | 0.91 | 0.18 | -0.48 | 0.63 | 0.62 | 1.34 |
| Highest level of education: | |  |  |  |  |  |  |
|  | A-Level | 0.69 | 0.07 | -3.85 | 0.00 | 0.57 | 0.84 |
|  | GCSE | 0.82 | 0.09 | -1.86 | 0.06 | 0.66 | 1.01 |
|  | Other | 0.61 | 0.13 | -2.37 | 0.02 | 0.41 | 0.92 |
|  | None | 0.36 | 0.13 | -2.86 | 0.00 | 0.18 | 0.72 |
| Self-reported financial conditions: bad | | 2.28 | 0.19 | 9.66 | 0.00 | 1.93 | 2.69 |
| Health condition: yes | | 2.14 | 0.18 | 9.34 | 0.00 | 1.83 | 2.52 |
| Company size: | |  |  |  |  |  |  |
|  | 25-199 | 0.88 | 0.08 | -1.37 | 0.17 | 0.73 | 1.06 |
|  | More than 200 | 0.96 | 0.09 | -0.44 | 0.66 | 0.79 | 1.16 |
| Wave Number | |  |  |  |  |  |  |
|  | Wave 10 | 0.92 | 0.09 | -0.83 | 0.41 | 0.76 | 1.12 |
|  | Wave 11 | 1.33 | 0.14 | 2.68 | 0.01 | 1.08 | 1.65 |
|  | COVID-19 Sweeps |  |  |  |  |  |  |
|  | Apr-20 | 2.25 | 0.25 | 7.21 | 0.00 | 1.80 | 2.80 |
|  | May-20 | 1.84 | 0.21 | 5.29 | 0.00 | 1.47 | 2.31 |
|  | Jun-20 | 1.69 | 0.21 | 4.30 | 0.00 | 1.33 | 2.14 |
|  | Jul-20 | 1.01 | 0.12 | 0.09 | 0.93 | 0.81 | 1.26 |
|  | Sep-20 | 0.95 | 0.12 | -0.38 | 0.70 | 0.74 | 1.22 |
|  | Nov-20 | 1.89 | 0.24 | 4.97 | 0.00 | 1.47 | 2.42 |
|  | Jan-21 | 1.63 | 0.20 | 4.03 | 0.00 | 1.28 | 2.06 |
|  | Mar-21 | 1.27 | 0.16 | 1.97 | 0.05 | 1.00 | 1.62 |
|  | Sep-21 | 1.00 | (omitted) |  |  |  |  |
| Constant | | 0.02 | 0.01 | -7.07 | 0.00 | 0.01 | 0.07 |

Note: Observations=39,190; Groups=4,405 (Minimum=1; Average= 7.8; Maximum=12).

#### Union presence – adjusted model (2-way interaction), excluding public workplaces

|  |  | **Adjusted model (weighted)** | | | | | |
| --- | --- | --- | --- | --- | --- | --- | --- |
|  |  | **OR** | **std. err.** | **z** | **P>z** | **[95% conf.** | **interval]** |
| sex |  | 2.29 | 0.20 | 9.73 | 0.00 | 1.94 | 2.71 |
| age |  | 1.02 | 0.02 | 0.78 | 0.44 | 0.97 | 1.07 |
| age_square | | 1.00 | 0.00 | -1.96 | 0.05 | 1.00 | 1.00 |
| Ethnicity | | 0.93 | 0.15 | -0.43 | 0.67 | 0.67 | 1.29 |
| Pre-Post Pandemic | | 1.55 | 0.22 | 3.13 | 0.00 | 1.18 | 2.03 |
| Union presence: yes | | 1.09 | 0.13 | 0.72 | 0.47 | 0.87 | 1.36 |
| Pre-Post Pandemic # Union presence: yes | | 0.85 | 0.11 | -1.25 | 0.21 | 0.65 | 1.10 |
| Industry: | |  |  |  |  |  |  |
|  | Mining, Energy and Water Supply | 1.36 | 0.36 | 1.16 | 0.25 | 0.81 | 2.30 |
|  | Manufacturing | 0.76 | 0.16 | -1.28 | 0.20 | 0.49 | 1.16 |
|  | Construction | 0.76 | 0.19 | -1.09 | 0.27 | 0.46 | 1.24 |
|  | Wholesale and Retail Trade Motor Repair | 0.99 | 0.21 | -0.07 | 0.94 | 0.65 | 1.49 |
|  | Transportation and Storage | 0.83 | 0.20 | -0.75 | 0.46 | 0.52 | 1.34 |
|  | Information and Communication | 0.89 | 0.21 | -0.52 | 0.60 | 0.56 | 1.40 |
|  | Financial and Insurance Activities | 0.72 | 0.18 | -1.35 | 0.18 | 0.44 | 1.16 |
|  | Real Estate Activities | 1.29 | 0.38 | 0.85 | 0.39 | 0.72 | 2.32 |
|  | Professional Scientific and Technical | 0.88 | 0.19 | -0.59 | 0.55 | 0.58 | 1.34 |
|  | Administrative and Support Services | 0.73 | 0.19 | -1.23 | 0.22 | 0.44 | 1.21 |
|  | Public Administration and Defence | 1.16 | 0.38 | 0.44 | 0.66 | 0.61 | 2.21 |
|  | Education | 0.91 | 0.20 | -0.40 | 0.69 | 0.59 | 1.42 |
|  | Human Health and Social Work Activities | 0.88 | 0.19 | -0.60 | 0.55 | 0.57 | 1.35 |
|  | Other Services | 0.75 | 0.16 | -1.30 | 0.19 | 0.49 | 1.15 |
| Country: | |  |  |  |  |  |  |
|  | Wales | 1.30 | 0.23 | 1.49 | 0.14 | 0.92 | 1.83 |
|  | Scotland | 0.90 | 0.15 | -0.61 | 0.54 | 0.65 | 1.26 |
|  | Northern Ireland | 0.79 | 0.16 | -1.16 | 0.25 | 0.52 | 1.18 |
| Highest level of education: | |  |  |  |  |  |  |
|  | A-Level | 0.72 | 0.07 | -3.30 | 0.00 | 0.59 | 0.87 |
|  | GCSE | 0.75 | 0.08 | -2.59 | 0.01 | 0.60 | 0.93 |
|  | Other | 0.61 | 0.13 | -2.38 | 0.02 | 0.40 | 0.92 |
|  | None | 0.46 | 0.17 | -2.14 | 0.03 | 0.23 | 0.94 |
| Self-reported financial conditions: bad | | 2.57 | 0.23 | 10.35 | 0.00 | 2.15 | 3.07 |
| Health condition: yes | | 2.28 | 0.20 | 9.22 | 0.00 | 1.92 | 2.72 |
| Company size: | |  |  |  |  |  |  |
|  | 25-199 | 0.85 | 0.09 | -1.60 | 0.11 | 0.69 | 1.04 |
|  | More than 200 | 0.88 | 0.09 | -1.25 | 0.21 | 0.72 | 1.07 |
| Wave Number | |  |  |  |  |  |  |
|  | Wave 10 | 0.89 | 0.09 | -1.15 | 0.25 | 0.72 | 1.09 |
|  | Wave 11 | 1.31 | 0.15 | 2.46 | 0.01 | 1.06 | 1.63 |
|  | COVID-19 Sweeps |  |  |  |  |  |  |
|  | Apr-20 | 2.17 | 0.27 | 6.16 | 0.00 | 1.70 | 2.78 |
|  | May-20 | 1.72 | 0.22 | 4.35 | 0.00 | 1.35 | 2.20 |
|  | Jun-20 | 1.53 | 0.20 | 3.19 | 0.00 | 1.18 | 1.98 |
|  | Jul-20 | 0.97 | 0.12 | -0.27 | 0.79 | 0.75 | 1.24 |
|  | Sep-20 | 1.01 | 0.14 | 0.11 | 0.92 | 0.78 | 1.32 |
|  | Nov-20 | 1.80 | 0.26 | 4.04 | 0.00 | 1.35 | 2.38 |
|  | Jan-21 | 1.58 | 0.21 | 3.36 | 0.00 | 1.21 | 2.06 |
|  | Mar-21 | 1.32 | 0.18 | 2.08 | 0.04 | 1.02 | 1.72 |
|  | Sep-21 | 1.00 | (omitted) |  |  |  |  |
| Constant |  | 0.03 | 0.02 | -6.31 | 0.00 | 0.01 | 0.08 |

Note: Observations=29,513; Groups=4,116 (Minimum=1; Average= 7.2; Maximum=12).

#### Union presence – adjusted model (3-way interaction), excluding public workplaces

|  |  | **Adjusted model (weighted) -- three-ways interaction** | | | | | |
| --- | --- | --- | --- | --- | --- | --- | --- |
|  |  | **OR** | **std. err.** | **z** | **P>z** | **[95% conf.** | **interval]** |
| Sex |  | 2.28 | 0.20 | 9.58 | 0.00 | 1.93 | 2.70 |
| Age | | 1.02 | 0.02 | 0.86 | 0.39 | 0.97 | 1.07 |
| Age square | | 1.00 | 0.00 | -2.05 | 0.04 | 1.00 | 1.00 |
| Ethniciy | | 0.93 | 0.16 | -0.42 | 0.68 | 0.67 | 1.29 |
| Pre-Post Pandemic # Industry: |  |  |  |  |  |  |  |
|  | Pre-Pandemic-#Mining, Energy and Water Supply | 3.31 | 1.95 | 2.04 | 0.04 | 1.05 | 10.50 |
|  | Post-Pandemic-#Mining, Energy and Water Supply | 2.51 | 0.86 | 2.70 | 0.01 | 1.29 | 4.89 |
|  | Pre-Pandemic-#Manufacturing | 0.86 | 0.22 | -0.60 | 0.55 | 0.52 | 1.42 |
|  | Post-Pandemic-#Manufacturing | 1.49 | 0.41 | 1.48 | 0.14 | 0.88 | 2.54 |
|  | Pre-Pandemic-#Construction | 0.70 | 0.23 | -1.08 | 0.28 | 0.36 | 1.34 |
|  | Post-Pandemic-#Construction | 1.27 | 0.37 | 0.81 | 0.42 | 0.72 | 2.24 |
|  | Pre-Pandemic-#Wholesale and Retail Trade Motor Repair | 1.15 | 0.31 | 0.52 | 0.60 | 0.68 | 1.96 |
|  | Post-Pandemic-#Wholesale and Retail Trade Motor Repair | 1.80 | 0.51 | 2.09 | 0.04 | 1.04 | 3.12 |
|  | Pre-Pandemic-#Transportation and Storage | 1.16 | 0.44 | 0.39 | 0.70 | 0.55 | 2.44 |
|  | Post-Pandemic-#Transportation and Storage | 1.06 | 0.37 | 0.17 | 0.87 | 0.53 | 2.10 |
|  | Pre-Pandemic-#Accommodation and Food Services | 1.56 | 0.53 | 1.31 | 0.19 | 0.80 | 3.03 |
|  | Post-Pandemic-#Accommodation and Food Services | 1.84 | 0.60 | 1.85 | 0.06 | 0.96 | 3.50 |
|  | Pre-Pandemic-#Information and Communication | 0.80 | 0.30 | -0.60 | 0.55 | 0.38 | 1.67 |
|  | Post-Pandemic-#Information and Communication | 1.81 | 0.47 | 2.26 | 0.02 | 1.08 | 3.02 |
|  | Pre-Pandemic-#Financial and Insurance Activities | 1.43 | 0.45 | 1.12 | 0.26 | 0.76 | 2.67 |
|  | Post-Pandemic-#Financial and Insurance Activities | 1.18 | 0.34 | 0.59 | 0.56 | 0.68 | 2.06 |
|  | Pre-Pandemic-#Real Estate Activities | 1.08 | 0.50 | 0.18 | 0.86 | 0.44 | 2.66 |
|  | Post-Pandemic-#Real Estate Activities | 1.81 | 0.78 | 1.39 | 0.17 | 0.78 | 4.21 |
|  | Pre-Pandemic-#Professional Scientific and Technical | 0.86 | 0.21 | -0.60 | 0.55 | 0.53 | 1.40 |
|  | Post-Pandemic-#Professional Scientific and Technical | 1.53 | 0.38 | 1.74 | 0.08 | 0.95 | 2.49 |
|  | Pre-Pandemic-#Administrative and Support Services | 0.67 | 0.25 | -1.10 | 0.27 | 0.32 | 1.37 |
|  | Post-Pandemic-#Administrative and Support Services | 1.42 | 0.43 | 1.15 | 0.25 | 0.78 | 2.58 |
|  | Pre-Pandemic-#Public Administration and Defence | 1.98 | 1.75 | 0.77 | 0.44 | 0.35 | 11.23 |
|  | Post-Pandemic-#Public Administration and Defence | 1.15 | 0.52 | 0.31 | 0.76 | 0.48 | 2.77 |
|  | Pre-Pandemic-#Education | 0.57 | 0.27 | -1.19 | 0.23 | 0.23 | 1.44 |
|  | Post-Pandemic-#Education | 1.55 | 0.48 | 1.41 | 0.16 | 0.84 | 2.85 |
|  | Post-Pandemic-#Human Health and Social Work Activities | 1.53 | 0.35 | 1.85 | 0.07 | 0.97 | 2.41 |
|  | Pre-Pandemic-#Other Services | 1.21 | 0.41 | 0.56 | 0.58 | 0.62 | 2.34 |
|  | Post-Pandemic-#Other Services | 1.45 | 0.36 | 1.50 | 0.13 | 0.89 | 2.34 |
| Union presence: yes | | 1.18 | 0.37 | 0.52 | 0.61 | 0.63 | 2.19 |
| Pre-Post Pandemic # Industry # Union presence | |  |  |  |  |  |  |
|  | Pre-Pandemic-#Mining, Energy and Water Supply#Yes | 0.36 | 0.29 | -1.26 | 0.21 | 0.07 | 1.77 |
|  | Post-Pandemic-#Mining, Energy and Water Supply#Yes | 0.57 | 0.30 | -1.07 | 0.29 | 0.21 | 1.59 |
|  | Pre-Pandemic-#Manufacturing#Yes | 0.62 | 0.29 | -1.01 | 0.31 | 0.25 | 1.56 |
|  | Post-Pandemic-#Manufacturing#Yes | 0.65 | 0.29 | -0.98 | 0.33 | 0.27 | 1.55 |
|  | Pre-Pandemic-#Construction#Yes | 2.13 | 1.45 | 1.11 | 0.27 | 0.56 | 8.13 |
|  | Post-Pandemic-#Construction#Yes | 1.11 | 0.58 | 0.20 | 0.85 | 0.40 | 3.09 |
|  | Pre-Pandemic-#Wholesale and Retail Trade Motor Repair#Yes | 0.92 | 0.40 | -0.20 | 0.84 | 0.39 | 2.14 |
|  | Post-Pandemic-#Wholesale and Retail Trade Motor Repair#Yes | 0.70 | 0.29 | -0.87 | 0.39 | 0.31 | 1.57 |
|  | Pre-Pandemic-#Transportation and Storage#Yes | 0.68 | 0.36 | -0.73 | 0.46 | 0.24 | 1.92 |
|  | Post-Pandemic-#Transportation and Storage#Yes | 1.30 | 0.64 | 0.54 | 0.59 | 0.50 | 3.41 |
|  | Pre-Pandemic-#Accommodation and Food Services#Yes | 0.24 | 0.19 | -1.77 | 0.08 | 0.05 | 1.16 |
|  | Post-Pandemic-#Accommodation and Food Services#Yes | 0.42 | 0.26 | -1.43 | 0.15 | 0.13 | 1.38 |
|  | Pre-Pandemic-#Information and Communication#Yes | 1.98 | 1.19 | 1.13 | 0.26 | 0.60 | 6.46 |
|  | Post-Pandemic-#Information and Communication#Yes | 0.45 | 0.24 | -1.47 | 0.14 | 0.15 | 1.30 |
|  | Pre-Pandemic-#Financial and Insurance Activities#Yes | 0.54 | 0.29 | -1.17 | 0.24 | 0.19 | 1.53 |
|  | Post-Pandemic-#Financial and Insurance Activities#Yes | 0.67 | 0.31 | -0.86 | 0.39 | 0.27 | 1.66 |
|  | Pre-Pandemic-#Real Estate Activities#Yes | 4.08 | 3.05 | 1.88 | 0.06 | 0.94 | 17.65 |
|  | Post-Pandemic-#Real Estate Activities#Yes | 0.78 | 0.59 | -0.33 | 0.74 | 0.17 | 3.45 |
|  | Pre-Pandemic-#Professional Scientific and Technical#Yes | 1.00 | 0.50 | 0.00 | 1.00 | 0.37 | 2.68 |
|  | Post-Pandemic-#Professional Scientific and Technical#Yes | 1.25 | 0.54 | 0.52 | 0.60 | 0.54 | 2.92 |
|  | Pre-Pandemic-#Administrative and Support Services#Yes | 1.00 | 0.76 | 0.00 | 1.00 | 0.23 | 4.40 |
|  | Post-Pandemic-#Administrative and Support Services#Yes | 0.71 | 0.43 | -0.56 | 0.57 | 0.22 | 2.30 |
|  | Pre-Pandemic-#Public Administration and Defence#Yes | 0.93 | 0.94 | -0.07 | 0.94 | 0.13 | 6.73 |
|  | Post-Pandemic-#Public Administration and Defence#Yes | 1.91 | 1.14 | 1.08 | 0.28 | 0.59 | 6.17 |
|  | Pre-Pandemic-#Education#Yes | 1.53 | 0.88 | 0.75 | 0.45 | 0.50 | 4.69 |
|  | Post-Pandemic-#Education#Yes | 1.20 | 0.53 | 0.41 | 0.68 | 0.50 | 2.85 |
|  | Post-Pandemic-#Human Health and Social Work Activities#Yes | 0.85 | 0.36 | -0.38 | 0.71 | 0.38 | 1.93 |
|  | Pre-Pandemic-#Other Services #Yes | 0.56 | 0.37 | -0.88 | 0.38 | 0.15 | 2.04 |
|  | Post-Pandemic-#Other Services #Yes | 0.52 | 0.23 | -1.48 | 0.14 | 0.22 | 1.24 |
| Country: | |  |  |  |  |  |  |
|  | Wales | 1.31 | 0.23 | 1.54 | 0.12 | 0.93 | 1.86 |
|  | Scotland | 0.90 | 0.15 | -0.63 | 0.53 | 0.64 | 1.26 |
|  | Northern Ireland | 0.79 | 0.17 | -1.14 | 0.26 | 0.52 | 1.19 |
| Highest level of education: | |  |  |  |  |  |  |
|  | A-Level | 0.72 | 0.07 | -3.16 | 0.00 | 0.59 | 0.88 |
|  | GCSE | 0.76 | 0.09 | -2.43 | 0.02 | 0.61 | 0.95 |
|  | Other | 0.62 | 0.13 | -2.26 | 0.02 | 0.41 | 0.94 |
|  | None | 0.49 | 0.18 | -1.97 | 0.05 | 0.24 | 1.00 |
| Self-reported financial conditions: bad | | 2.55 | 0.23 | 10.20 | 0.00 | 2.13 | 3.05 |
| Health condition: yes | | 2.29 | 0.21 | 9.22 | 0.00 | 1.92 | 2.73 |
| Company size: | |  |  |  |  |  |  |
|  | 25-199 | 0.85 | 0.09 | -1.62 | 0.11 | 0.69 | 1.04 |
|  | More than 200 | 0.88 | 0.09 | -1.23 | 0.22 | 0.72 | 1.08 |
| Wave Number | |  |  |  |  |  |  |
|  | Wave 10 | 0.88 | 0.09 | -1.19 | 0.23 | 0.72 | 1.08 |
|  | Wave 11 | 1.32 | 0.15 | 2.55 | 0.01 | 1.07 | 1.64 |
|  | COVID-19 Sweeps |  |  |  |  |  |  |
|  | Apr-20 | 2.17 | 0.27 | 6.12 | 0.00 | 1.69 | 2.77 |
|  | May-20 | 1.71 | 0.21 | 4.30 | 0.00 | 1.34 | 2.19 |
|  | Jun-20 | 1.53 | 0.20 | 3.20 | 0.00 | 1.18 | 1.99 |
|  | Jul-20 | 0.96 | 0.12 | -0.31 | 0.76 | 0.75 | 1.23 |
|  | Sep-20 | 1.01 | 0.14 | 0.06 | 0.95 | 0.77 | 1.32 |
|  | Nov-20 | 1.79 | 0.26 | 4.01 | 0.00 | 1.35 | 2.38 |
|  | Jan-21 | 1.57 | 0.21 | 3.32 | 0.00 | 1.20 | 2.04 |
|  | Mar-21 | 1.32 | 0.18 | 2.08 | 0.04 | 1.02 | 1.72 |
|  | Sep-21 | 1.00 | (omitted) |  |  |  |  |
| Constant | | 0.02 | 0.01 | -6.42 | 0.00 | 0.01 | 0.07 |

Note: Observations=29,513; Groups=4,116 (Minimum=1; Average= 7.2; Maximum=12).

#### Union presence – adjusted model (2-way interaction), GHQ-36

|  |  | **Adjusted model (weighted)** | | | | | |
| --- | --- | --- | --- | --- | --- | --- | --- |
|  |  | **OR** | **std. err.** | **z** | **P>z** | **[95% conf.** | **interval]** |
| sex |  | 2.16 | 0.15 | 11.42 | 0.00 | 1.89 | 2.47 |
| age |  | 1.01 | 0.02 | 0.32 | 0.75 | 0.97 | 1.04 |
| age_square | | 1.00 | 1.00 | 0.00 | -1.57 | 0.12 | 1.00 |
| Ethnicity | | 0.88 | 0.90 | 0.11 | -0.85 | 0.40 | 0.72 |
| Pre-Post Pandemic | | 1.53 | 1.45 | 0.16 | 3.34 | 0.00 | 1.17 |
| Union presence: yes | | 0.97 | 1.11 | 0.10 | 1.18 | 0.24 | 0.93 |
| Pre-Post Pandemic # Union presence: yes | | 0.94 | 0.89 | 0.09 | -1.23 | 0.22 | 0.73 |
| Industry: | |  |  |  |  |  |  |
|  | Mining, Energy and Water Supply | 1.13 | 0.29 | 0.47 | 0.64 | 0.68 | 1.86 |
|  | Manufacturing | 0.66 | 0.13 | -2.16 | 0.03 | 0.45 | 0.96 |
|  | Construction | 0.68 | 0.15 | -1.75 | 0.08 | 0.44 | 1.05 |
|  | Wholesale and Retail Trade Motor Repair | 0.85 | 0.16 | -0.90 | 0.37 | 0.59 | 1.22 |
|  | Transportation and Storage | 0.68 | 0.15 | -1.80 | 0.07 | 0.44 | 1.04 |
|  | Information and Communication | 0.85 | 0.18 | -0.77 | 0.44 | 0.57 | 1.28 |
|  | Financial and Insurance Activities | 0.64 | 0.14 | -2.03 | 0.04 | 0.42 | 0.98 |
|  | Real Estate Activities | 0.96 | 0.27 | -0.16 | 0.88 | 0.55 | 1.65 |
|  | Professional Scientific and Technical | 0.79 | 0.15 | -1.24 | 0.21 | 0.55 | 1.14 |
|  | Administrative and Support Services | 0.62 | 0.13 | -2.32 | 0.02 | 0.41 | 0.93 |
|  | Public Administration and Defence | 0.75 | 0.14 | -1.54 | 0.12 | 0.52 | 1.08 |
|  | Education | 0.78 | 0.14 | -1.36 | 0.17 | 0.55 | 1.11 |
|  | Human Health and Social Work Activities | 0.81 | 0.14 | -1.21 | 0.23 | 0.57 | 1.14 |
|  | Other Services | 0.67 | 0.13 | -2.12 | 0.03 | 0.47 | 0.97 |
| Country: | |  |  |  |  |  |  |
|  | Wales | 1.24 | 0.16 | 1.70 | 0.09 | 0.97 | 1.59 |
|  | Scotland | 0.98 | 0.11 | -0.17 | 0.86 | 0.79 | 1.22 |
|  | Northern Ireland | 0.87 | 0.13 | -0.92 | 0.36 | 0.65 | 1.17 |
| Highest level of education: | |  |  |  |  |  |  |
|  | A-Level | 1.24 | 0.16 | 1.70 | 0.09 | 0.97 | 1.59 |
|  | GCSE | 0.98 | 0.11 | -0.17 | 0.86 | 0.79 | 1.22 |
|  | Other | 0.87 | 0.13 | -0.92 | 0.36 | 0.65 | 1.17 |
|  | None | 0.37 | 0.12 | -3.19 | 0.00 | 0.20 | 0.68 |
| Self-reported financial conditions: bad | | 1.24 | 0.16 | 1.70 | 0.09 | 0.97 | 1.59 |
| Health condition: yes | | 0.98 | 0.11 | -0.17 | 0.86 | 0.79 | 1.22 |
| Company size: | |  |  |  |  |  |  |
|  | 25-199 | 0.89 | 0.07 | -1.46 | 0.14 | 0.76 | 1.04 |
|  | More than 200 | 0.95 | 0.07 | -0.64 | 0.52 | 0.81 | 1.11 |
| Wave Number | |  |  |  |  |  |  |
|  | Wave 10 | 0.92 | 0.09 | -0.80 | 0.42 | 0.76 | 1.12 |
|  | Wave 11 | 1.31 | 0.14 | 2.53 | 0.01 | 1.06 | 1.63 |
|  | COVID-19 Sweeps |  |  |  |  |  |  |
|  | Apr-20 | 0.91 | 0.07 | -1.23 | 0.22 | 0.78 | 1.06 |
|  | May-20 | 1.22 | 0.11 | 2.32 | 0.02 | 1.03 | 1.45 |
|  | Jun-20 | 2.29 | 0.20 | 9.22 | 0.00 | 1.92 | 2.73 |
|  | Jul-20 | 1.81 | 0.17 | 6.47 | 0.00 | 1.51 | 2.16 |
|  | Sep-20 | 1.62 | 0.16 | 4.98 | 0.00 | 1.34 | 1.96 |
|  | Nov-20 | 0.96 | 0.09 | -0.44 | 0.66 | 0.80 | 1.15 |
|  | Jan-21 | 1.04 | 0.10 | 0.42 | 0.68 | 0.86 | 1.26 |
|  | Mar-21 | 1.81 | 0.18 | 5.82 | 0.00 | 1.48 | 2.21 |
|  | Sep-21 | 1.00 | (omitted) |  |  |  |  |
| Constant |  | 0.04 | 0.02 | -7.06 | 0.00 | 0.02 | 0.10 |

Note: Observations=49,915; Groups=5,988 (Minimum=1; Average= 8.3; Maximum=12).

#### Union membership – adjusted model (2-way interaction), GHQ-36

|  |  | **Adjusted model (weighted)** | | | | | |
| --- | --- | --- | --- | --- | --- | --- | --- |
|  |  | **OR** | **std. err.** | **z** | **P>z** | **[95% conf.** | **interval]** |
| sex |  | 2.08 | 0.20 | 7.81 | 0.00 | 1.73 | 2.50 |
| age |  | 1.02 | 0.03 | 0.77 | 0.44 | 0.97 | 1.08 |
| age_square | | 1.00 | 0.00 | -1.32 | 0.19 | 1.00 | 1.00 |
| Ethnicity | | 0.82 | 0.12 | -1.40 | 0.16 | 0.61 | 1.08 |
| Pre-Post Pandemic | | 1.40 | 0.19 | 2.43 | 0.02 | 1.07 | 1.83 |
| Union presence: yes | | 1.17 | 0.14 | 1.37 | 0.17 | 0.93 | 1.47 |
| Pre-Post Pandemic # Union presence: yes | | 0.84 | 0.11 | -1.35 | 0.18 | 0.66 | 1.08 |
| Industry: | |  |  |  |  |  |  |
|  | Mining, Energy and Water Supply | 1.08 | 0.44 | 0.20 | 0.84 | 0.49 | 2.42 |
|  | Manufacturing | 0.62 | 0.20 | -1.46 | 0.14 | 0.33 | 1.18 |
|  | Construction | 1.08 | 0.42 | 0.21 | 0.84 | 0.51 | 2.32 |
|  | Wholesale and Retail Trade Motor Repair | 1.07 | 0.32 | 0.21 | 0.83 | 0.59 | 1.93 |
|  | Transportation and Storage | 1.02 | 0.31 | 0.08 | 0.94 | 0.56 | 1.86 |
|  | Information and Communication | 1.18 | 0.40 | 0.49 | 0.63 | 0.61 | 2.30 |
|  | Financial and Insurance Activities | 0.63 | 0.21 | -1.37 | 0.17 | 0.32 | 1.22 |
|  | Real Estate Activities | 1.35 | 0.51 | 0.80 | 0.43 | 0.65 | 2.82 |
|  | Professional Scientific and Technical | 1.16 | 0.34 | 0.51 | 0.61 | 0.65 | 2.06 |
|  | Administrative and Support Services | 0.82 | 0.26 | -0.63 | 0.53 | 0.44 | 1.52 |
|  | Public Administration and Defence | 1.00 | 0.27 | 0.00 | 1.00 | 0.58 | 1.71 |
|  | Education | 1.05 | 0.28 | 0.19 | 0.85 | 0.62 | 1.77 |
|  | Human Health and Social Work Activities | 1.11 | 0.29 | 0.40 | 0.69 | 0.66 | 1.87 |
|  | Other Services | 0.80 | 0.23 | -0.78 | 0.44 | 0.45 | 1.41 |
| Country: | |  |  |  |  |  |  |
|  | Wales | 1.04 | 0.17 | 0.26 | 0.79 | 0.76 | 1.43 |
|  | Scotland | 0.94 | 0.12 | -0.49 | 0.62 | 0.73 | 1.21 |
|  | Northern Ireland | 0.89 | 0.16 | -0.66 | 0.51 | 0.63 | 1.26 |
| Highest level of education: | |  |  |  |  |  |  |
|  | A-Level | 0.75 | 0.08 | -2.66 | 0.01 | 0.61 | 0.93 |
|  | GCSE | 0.64 | 0.08 | -3.58 | 0.00 | 0.50 | 0.82 |
|  | Other | 0.45 | 0.11 | -3.34 | 0.00 | 0.28 | 0.72 |
|  | None | 0.42 | 0.18 | -2.00 | 0.05 | 0.18 | 0.98 |
| Self-reported financial conditions: bad | | 2.74 | 0.22 | 12.30 | 0.00 | 2.33 | 3.22 |
| Health condition: yes | | 1.94 | 0.17 | 7.57 | 0.00 | 1.63 | 2.30 |
| Company size: | |  |  |  |  |  |  |
|  | 25-199 | 0.95 | 0.11 | -0.45 | 0.65 | 0.75 | 1.19 |
|  | More than 200 | 1.05 | 0.12 | 0.48 | 0.63 | 0.85 | 1.31 |
| Wave Number | |  |  |  |  |  |  |
|  | Wave 10 | 0.89 | 0.09 | -1.12 | 0.26 | 0.72 | 1.09 |
|  | Wave 11 | 1.35 | 0.16 | 2.51 | 0.01 | 1.07 | 1.70 |
|  | COVID-19 Sweeps |  |  |  |  |  |  |
|  | Apr-20 | 2.57 | 0.28 | 8.63 | 0.00 | 2.07 | 3.18 |
|  | May-20 | 1.78 | 0.20 | 5.02 | 0.00 | 1.42 | 2.23 |
|  | Jun-20 | 1.55 | 0.19 | 3.55 | 0.00 | 1.22 | 1.98 |
|  | Jul-20 | 0.87 | 0.10 | -1.19 | 0.23 | 0.69 | 1.10 |
|  | Sep-20 | 1.13 | 0.13 | 0.99 | 0.32 | 0.89 | 1.42 |
|  | Nov-20 | 1.95 | 0.24 | 5.35 | 0.00 | 1.53 | 2.50 |
|  | Jan-21 | 2.23 | 0.25 | 7.02 | 0.00 | 1.78 | 2.79 |
|  | Mar-21 | 1.27 | 0.16 | 1.93 | 0.05 | 1.00 | 1.62 |
|  | Sep-21 | 1.00 | (omitted) |  |  |  |  |
| Constant |  | 0.02 | 0.01 | -5.65 | 0.00 | 0.01 | 0.08 |

Note: Observations=27,971; Groups=3,341 (Minimum=1; Average= 8.4; Maximum=12).
